# SARS-CoV-2 infection dynamics and genomic surveillance reveals early variant transmission in urban wastewater

**DOI:** 10.1101/2022.07.14.22277616

**Authors:** Sanjay Lamba, G Sutharsan, Namrta Daroch, Kiran Paul, Soumya Gopal Joshi, S Darshan, N Annamalai, S Vishwanath, Rakesh Mishra, Uma Ramakrishnan, Farah Ishtiaq

**Affiliations:** Tata Institute for Genetics and Society, GKVK Post, Bellary Road, Bangalore-560065, India; National Centre for Biological Sciences, TIFR, Bellary Road, Bangalore-560065, India; Biome Environmental Trust, Bangalore-560097, India

## Abstract

Environmental surveillance (ES) of a pathogen is crucial for understanding the community load of disease. As an early warning system, ES for SARS-CoV-2 has complemented routine diagnostic surveillance by capturing near real-time virus circulation at a population level. In this longitudinal study in 28 sewershed sites in Bangalore city, we quantified SARS-CoV-2 RNA to track infection dynamics and provide evidence of change in the relative abundance of emerging variants. We describe an early warning system using the exponentially weighted moving average control chart and demonstrate how SARS-CoV-2 RNA concentrations in wastewater correlated with clinically diagnosed new COVID-19 cases, with the trends appearing 8–14 days earlier in wastewater than in clinical data. This was further corroborated by showing that the estimated number of infections is strongly correlated with SARS-CoV-2 RNA copies detected in the wastewater. Using a deconvolution matrix, we detected emerging variants of concern up to two months earlier in wastewater samples. In addition, we found a huge diversity in variants detected in wastewater compared to clinical samples. Our study highlights that quantifying viral titres, correlating it with a known number of cases in the area, and combined with genomic surveillance helps in tracking VOCs over time and space, enabling timely and making informed policy decisions.

## Introduction

Globally, the COVID-19 pandemic (infectious pneumonia caused by Severe Acute Respiratory Syndrome Coronavirus 2: SARS-CoV-2) has brought Wastewater-based (sewage) epidemiology (WBE) to the forefront of the health surveillance system. Wastewater testing can provide a parallel and complementary snapshot of community health. WBE has become an integral component of environmental surveillance in more than 60 countries covering over 3000 sites^1,2^ providing near-real time information on health and community exposure to COVID-19^3^. Analysis of sewage identified evidence of SARS-CoV-2 RNA circulating 56 days in advance of the first clinically confirmed case in South America and 91 days in advance of the first clinically confirmed case in Brazil^4^, highlighting the crucial role of WBE in disease surveillance. Mostly, detection of SARS-CoV-2 in wastewater was correlated with local COVID-19 incidence, preceding the increase in clinical cases, locally by 1 to 3 weeks. Furthermore, WBE has also been used to detect regionally prevalent variants of SARS-CoV-2^5^. Given its potential to be a relatively widespread, economical, and rapid surveillance tool, WBE provides an excellent opportunity for a developing country like India, where issues such as unprecedented population growth (especially in urban centres), migrating populations, lack of public health systems and lack of integrated health surveillance are a reality.

WBE was first identified as an effective population wide monitoring technique over 40 years ago– with a high capacity for retrospective indication of poliovirus, norovirus, influenza, hepatitis and measles outbreaks^6^. Nonetheless, it has rarely been integrated into the health surveillance system in India except for environmental surveillance of poliovirus (Global polio eradication initiative. e.g.,^7^). Environmental surveillance played a crucial role in poliovirus eradication in India in 2012 and continues to provide vital support in documenting the absence of the polio virus in conjunction with active surveillance of acute flaccid paralysis (AFP) cases^7-9^.

In India, tracking of the COVID-19 pandemic relies heavily on testing symptomatic individuals for the presence of SARS-CoV-2 RNA and counting the positive tests over time. With high population density, many SARS-CoV-2 infected persons are likely to be asymptomatic or oligosymptomatic. They are generally not clinically tested, leading to underestimation of COVID-19 trends and prevalence. There is a need for new strategies to track the emergence and spread of SARS-CoV-2 variants. Currently, epidemiological surveillance relies on testing symptomatic cases using the respiratory tract as the principal site for virus replication.

However, the virus also replicates in the gastrointestinal tract leading to a high viral load in excreta^10^. The qPCR-based approach has revealed a strong correlation between SARS-CoV-2 incidence rates and the viral load in wastewater ^11-13^. With the continued emergence of new variants, we also need new approaches to identify the diversity between variants that might have escaped individual testing. Alternatively, emerging varianta in wastewater suggests that a significant proportion of individuals in the community are infected with it and thus, shedding the virus.

WBE is a powerful approach where epidemiological data must combine with genomic surveillance to gain insights into how the virus populations are changing, and how the virus is evolving. How long does it take for new variants to emerge and spread across cities (major hubs with international airports) and to rural or suburban areas? Wastewater samples are a mixed representative of local lineages circulating in the community. Monitoring SARS-CoV-2 lineages using wastewater has remained challenging due to low-quality reads, fragmented sequences, and the inability to estimate the relative lineage abundance based on patchy variant-defining mutations in a mixed community sample. Furthermore, SARS-CoV-2 lineages classification using pangolin^14^ and UShER^15^, designed for clinical samples often contain a single dominant variant and tend to underestimate the relative abundances of multiple SARS-CoV-2 lineages in samples with mixtures of viral genomes such as wastewater.

Using longitudinal testing of wastewater and quantitative epidemiological modelling, we aimed to investigate the relationship between wastewater SARS-CoV-2 concentrations, and COVID-19 cases. In addition, we used high throughput sequencing techniques, to quantify the prevalence and genetic diversity including the newly emerging SARS-CoV-2 variants in Bangalore city (12.9716° N, 77.5946° E, Karnataka, India). Wastewater samples from the inlet of 28 sewage treatment plants covering over 11 million people are being monitored for SARS-CoV-2 between August 2021-June 2022. We used next-generation sequencing of SARS-CoV-2 RNA and modelling of viral concentration from wastewater to explore the dynamics in diversity and abundance of SARS-CoV-2 lineages in Bangalore city. Our longitudinal study was initiated post second wave (Delta wave) from August 2021 through to the third wave (Omicron wave). Here we present results from January till June 2022 to show diversity in lineages of SARS-CoV-2 in wastewater and their comparison with the clinical data. This allows us to test the relationship between an increase in viral concentrations in wastewater with observed variant diversity across spatiotemporal scales. We hypothesize that high variant diversity is expected during a low viral load/low infection rate period. However, there should be a transition in the lineage of the dominant variant at the peak of infection period during high infection rates. In addition, we explore the emergence of new variants that would have escaped identification in clinical surveillance. Furthermore, we evaluate the lag between emerging new lineages in symptomatic individuals and wastewater samples representing community data.

## Results

### Longitudinal wastewater sampling can predict SARS-CoV-2 rise 1-2 weeks in advance

The normalised temporal trend of SARS-CoV-2 RNA in wastewater and new positive cases in Bangalore city are shown in Fig.2. Pearson’s correlation for citywide viral load showed a marginally stronger correlation with citywide COVID-19 cases with a time lag of 7-days than 4-days (R^2^=0.87, p=0.00001; Fig. 3 A & B). The sensitivity analysis showed that the estimated number of infections strongly correlates with the log SARS-CoV-2 RNA copies detected in the wastewater (Fig. 3. C & D).

**Figure 1.**
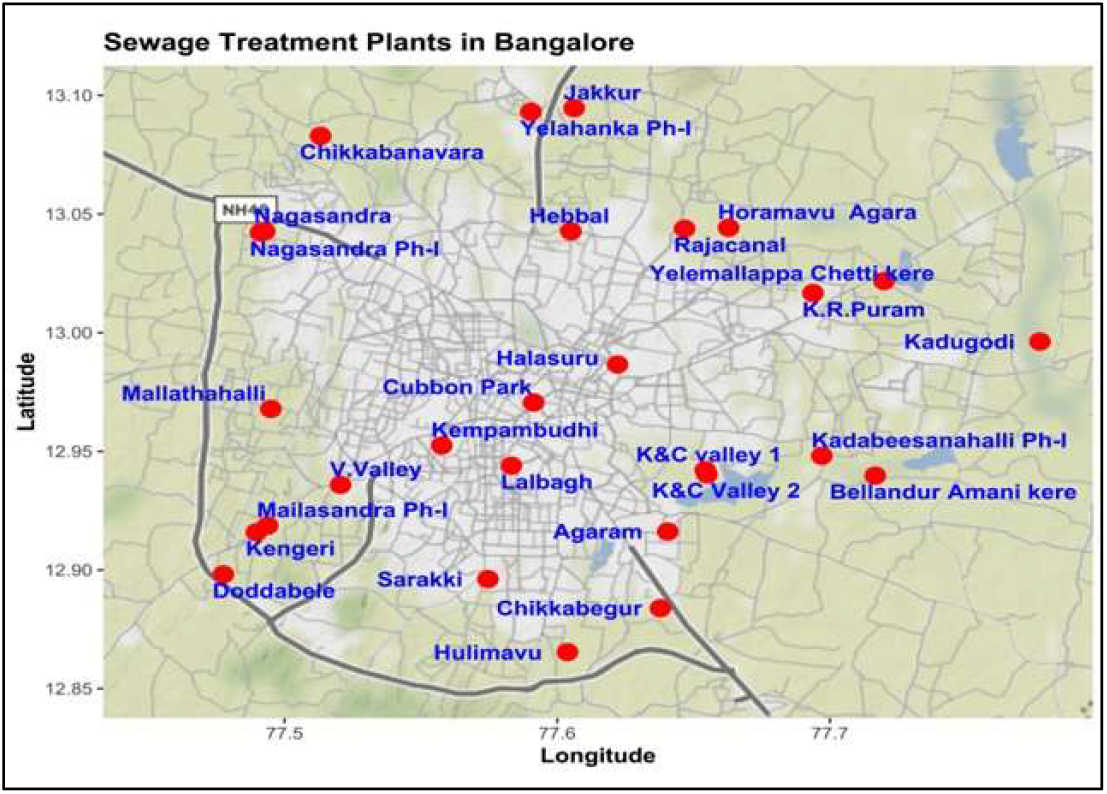
Location (red dots) of sewage treatment plants sampled once a week in Bangalore.

**Figure 2.**
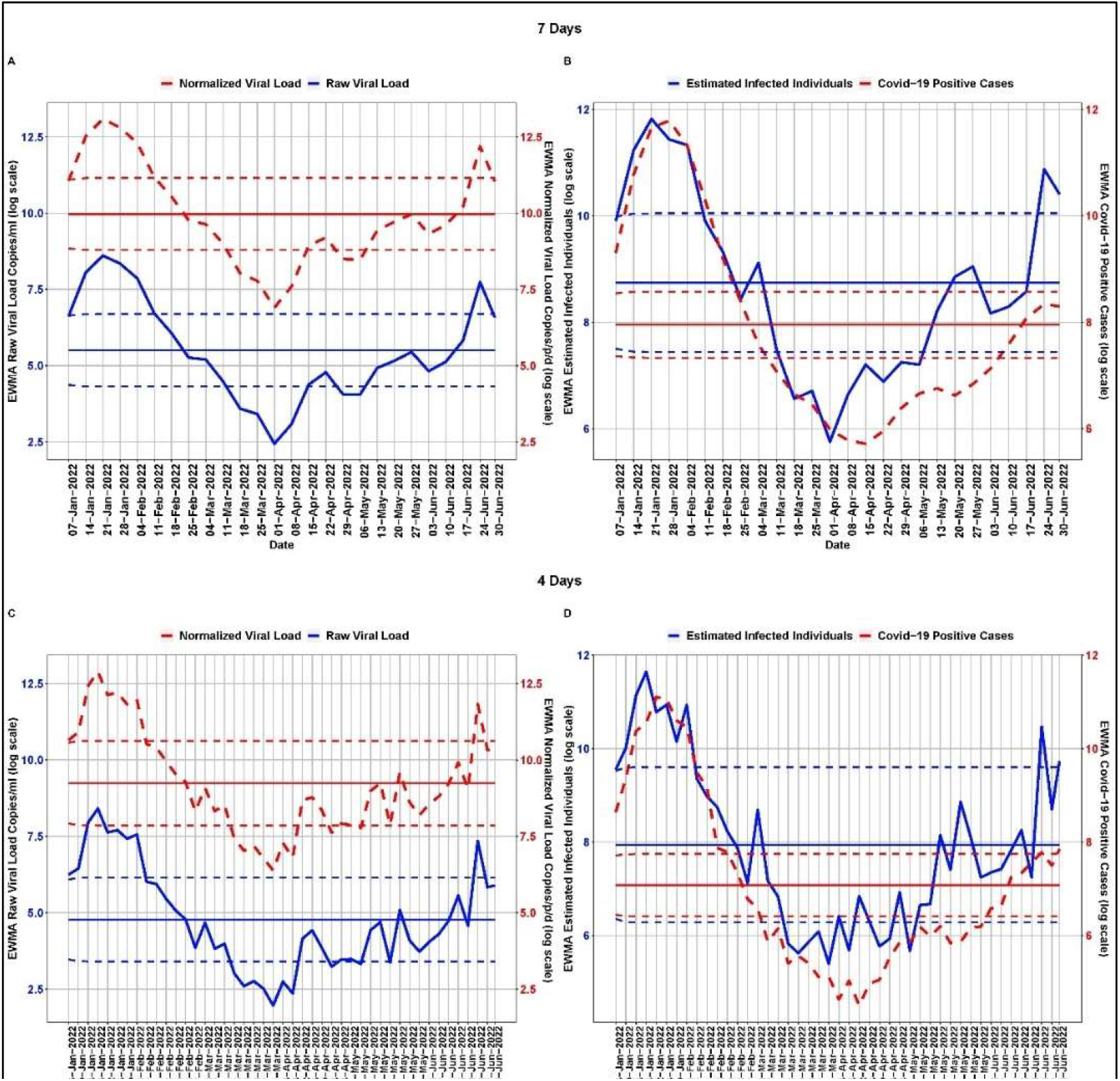
Temporal dynamics of normalized viral load in wastewater and COVID-19 cases in Bangalore. The dark centre red line shows the mean viral load, the dotted line above the mean is the upper control limit and the dotted line below the mean is the lower control limit. The 3^rd^ wave started in late December 2021 with peak in January and dropped down in February 2022.

**Figure 3.**
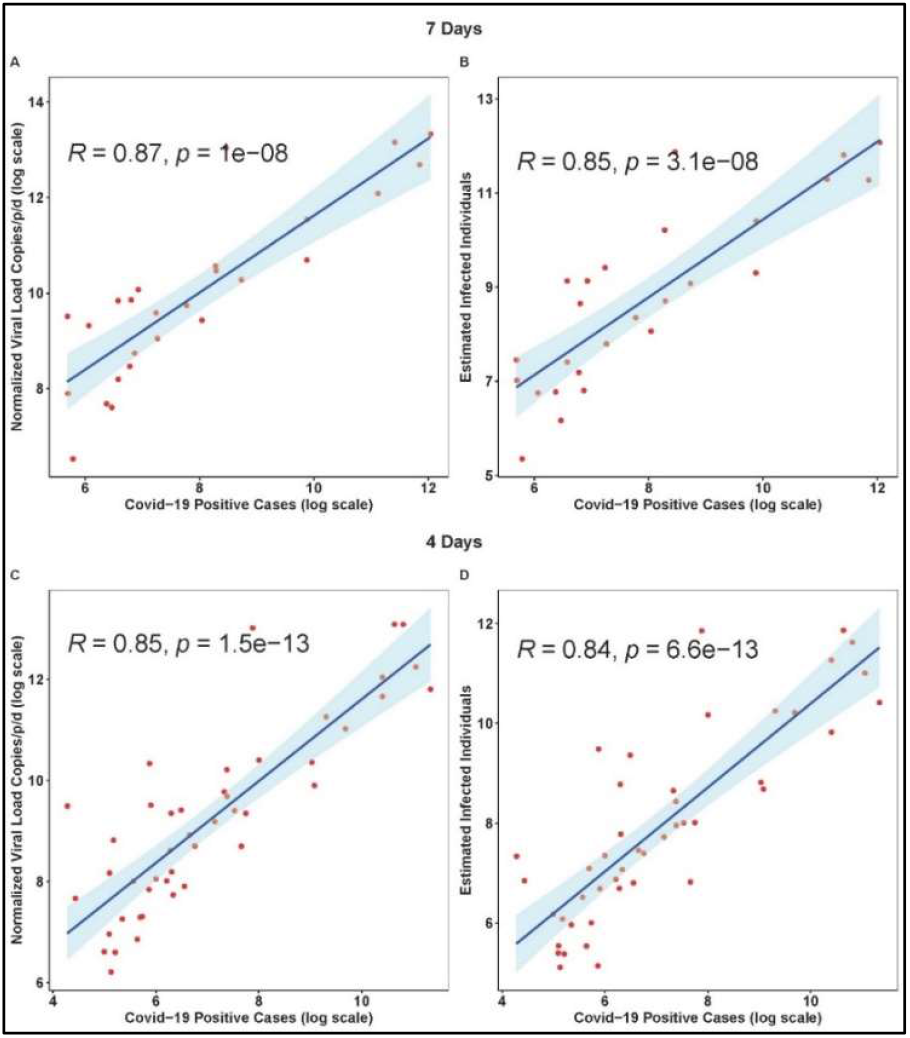
Viral concentrations correlate with daily new cases with a 4-d and 7-d time lag in Bangalore city. The Blue solid line is the linear regression fitting. Light blue area: 95% confidence interval from standard error of the fitting.

We used EWMA of RNA concentration in wastewater (smoothing parameter=0.7, *ρ* = 2) and new clinical cases (smoothing parameter=0.7, *ρ* = 2) to show the qualitative trend in the city. Evidently, there were two major outbreaks-January and June 2022. The EWMA captured the increased viral load and associated estimated infection during the peak of the SARS-CoV-2 wave in January 2022. The EWMA showed an increased viral trend in June as ‘red alert’ and associated estimated number of infections at least 14 days and 8 days in advance using time-lags of 7 days and 4-days, respectively (Table 1). SARS-CoV-2 was detected throughout the period with a low viral load of 6.38 copies/person/day (log scale) from 27^th^ March to 8^th^ April 2022. While the viral load pattern mirrored with the clinical data, COVID-19 positive cases appeared to remain underreported in the city. The estimated number of infected cases remained high indicating viral detection in wastewater in the early stage (below LCL) of the local outbreak (Fig. 2). Our EWMA analysis showed that using a time lag of 7 days, the increased estimated infected individuals trend reached the red alert (UCL ∼ 23106 cases/week) whereas COVID-19 cases were near the mean (threshold value ∼2847 cases/week). Using a time lag of 4 days, the estimated infected individuals reached the red alert (UCL ∼ 14803 cases / 4 days) whereas COVID-19 cases were near the mean and were rising to reach the red alert (UCL ∼2310 cases / 4 days).

**Table 1.**
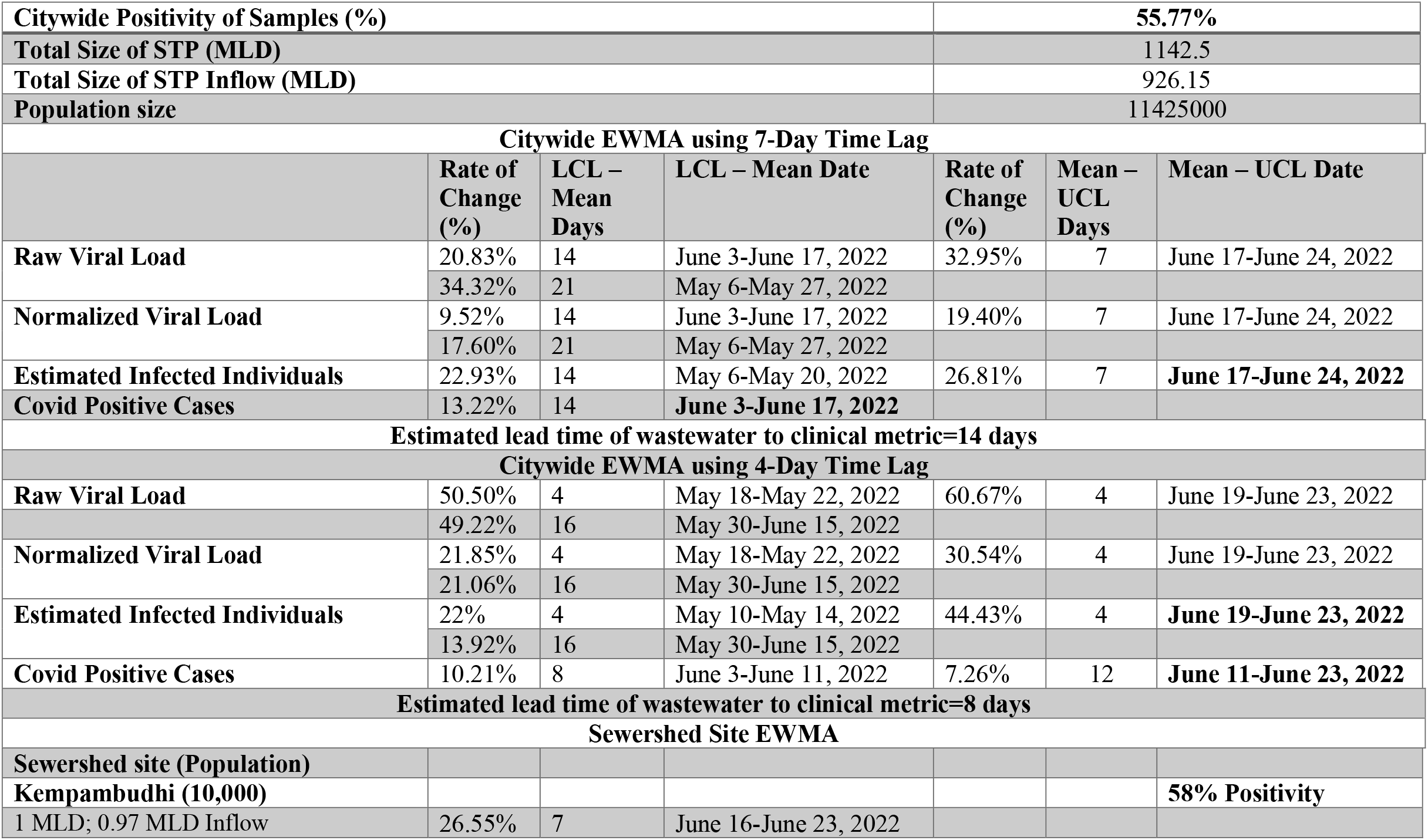
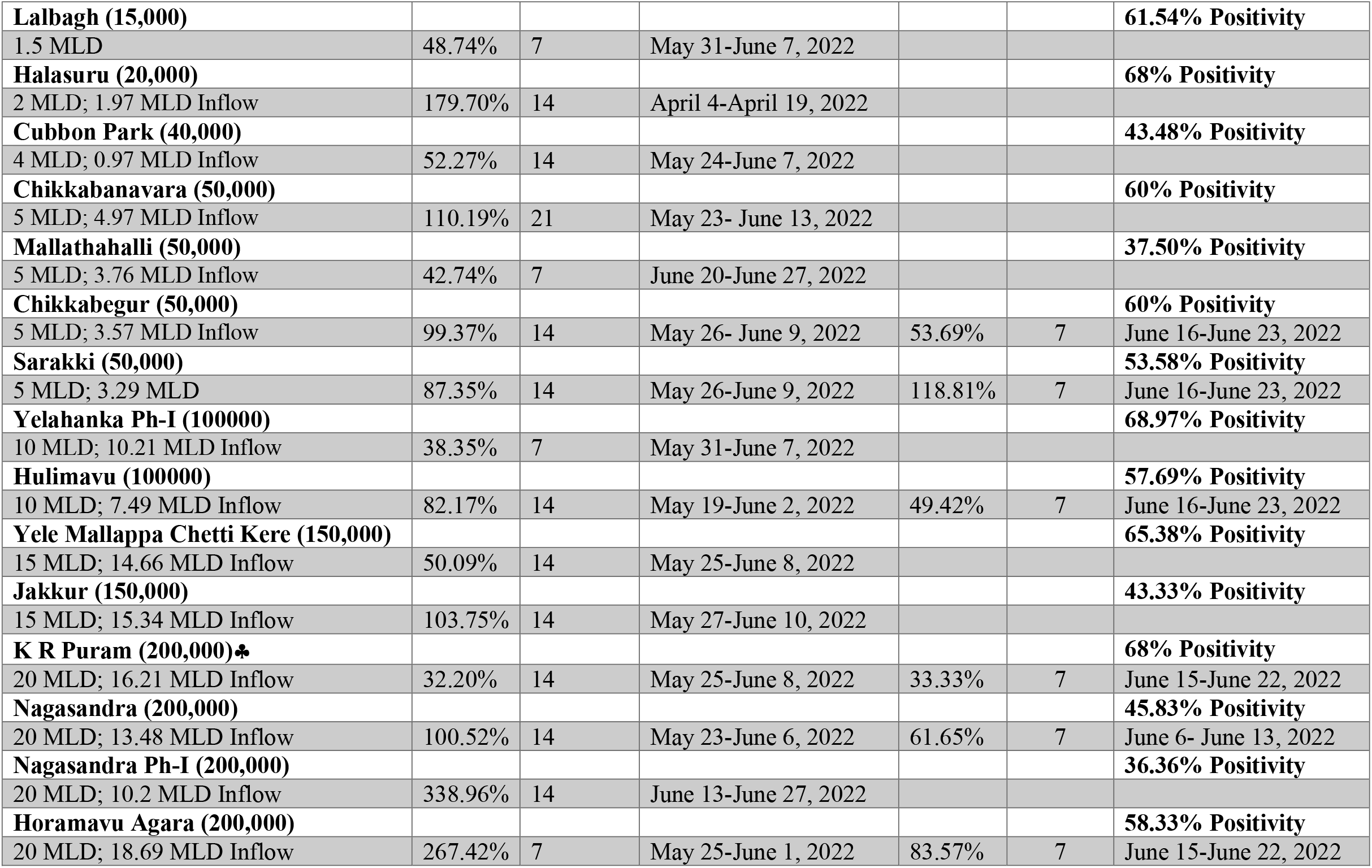
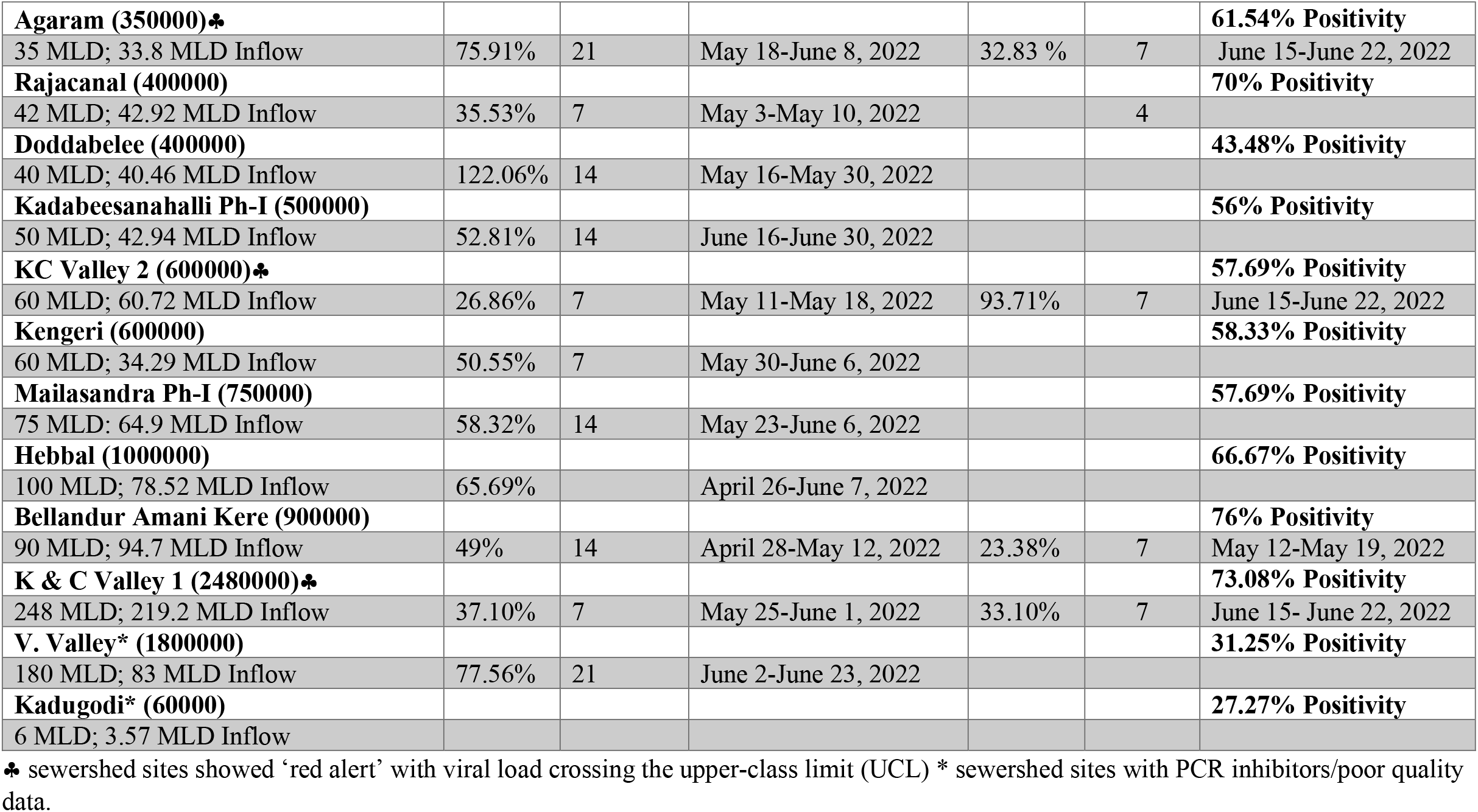
Lead time of normalized SARS-CoV-2 viral RNA signal weekly SARS-CoV-2 percent positivity.

|  |  |  |  |  |  |  |
| --- | --- | --- | --- | --- | --- | --- |
| Citywide Positivity of Samples (%) |  |  |  | 55.77% |  |  |
| Total Size of STP (MLD) |  |  |  | 1142.5 |  |  |
| Total Size of STP Inflow (MLD) |  |  |  | 926.15 |  |  |
| Population size |  |  |  | 11425000 |  |  |
| Citywide EWMA using 7-Day Time Lag |  |  |  |  |  |  |
|  | Rate of Change (%) | LCL – Mean Days | LCL – Mean Date | Rate of Change (%) | Mean – UCL Days | Mean – UCL Date |
| Raw Viral Load | 20.83% | 14 | June 3-June 17, 2022 | 32.95% | 7 | June 17-June 24, 2022 |
|  | 34.32% | 21 | May 6-May 27, 2022 |  |  |  |
| Normalized Viral Load | 9.52% | 14 | June 3-June 17, 2022 | 19.40% | 7 | June 17-June 24, 2022 |
|  | 17.60% | 21 | May 6-May 27, 2022 |  |  |  |
| Estimated Infected Individuals | 22.93% | 14 | May 6-May 20, 2022 | 26.81% | 7 | June 17-June 24, 2022 |
| Covid Positive Cases | 13.22% | 14 | June 3-June 17, 2022 |  |  |  |
| Estimated lead time of wastewater to clinical metric=14 days |  |  |  |  |  |  |
| Citywide EWMA using 4-Day Time Lag |  |  |  |  |  |  |
| Raw Viral Load | 50.50% | 4 | May 18-May 22, 2022 | 60.67% | 4 | June 19-June 23, 2022 |
|  | 49.22% | 16 | May 30-June 15, 2022 |  |  |  |
| Normalized Viral Load | 21.85% | 4 | May 18-May 22, 2022 | 30.54% | 4 | June 19-June 23, 2022 |
|  | 21.06% | 16 | May 30-June 15, 2022 |  |  |  |
| Estimated Infected Individuals | 22% | 4 | May 10-May 14, 2022 | 44.43% | 4 | June 19-June 23, 2022 |
|  | 13.92% | 16 | May 30-June 15, 2022 |  |  |  |
| Covid Positive Cases | 10.21% | 8 | June 3-June 11, 2022 | 7.26% | 12 | June 11-June 23, 2022 |
| Estimated lead time of wastewater to clinical metric=8 days |  |  |  |  |  |  |
| Sewershed Site EWMA |  |  |  |  |  |  |
| Sewershed site (Population) |  |  |  |  |  |  |
| Kempambudhi (10,000) |  |  |  |  |  | 58% Positivity |
| 1 MLD; 0.97 MLD Inflow | 26.55% | 7 | June 16-June 23, 2022 |  |  |  |
| **Lalbagh (15,000)** |  |  |  |  |  | **61.54% Positivity** |
| 1.5 MLD | 48.74% | 7 | May 31-June 7, 2022 |  |  |  |
| **Halasuru (20,000)** |  |  |  |  |  | **68% Positivity** |
| 2 MLD; 1.97 MLD Inflow | 179.70% | 14 | April 4-April 19, 2022 |  |  |  |
| **Cubbon Park (40,000)** |  |  |  |  |  | **43.48% Positivity** |
| 4 MLD; 0.97 MLD Inflow | 52.27% | 14 | May 24-June 7, 2022 |  |  |  |
| **Chikkabanavara (50,000)** |  |  |  |  |  | **60% Positivity** |
| 5 MLD; 4.97 MLD Inflow | 110.19% | 21 | May 23- June 13, 2022 |  |  |  |
| **Mallathahalli (50,000)** |  |  |  |  |  | **37.50% Positivity** |
| 5 MLD; 3.76 MLD Inflow | 42.74% | 7 | June 20-June 27, 2022 |  |  |  |
| **Chikkabegur (50,000)** |  |  |  |  |  | **60% Positivity** |
| 5 MLD; 3.57 MLD Inflow | 99.37% | 14 | May 26- June 9, 2022 | 53.69% | 7 | June 16-June 23, 2022 |
| **Sarakki (50,000)** |  |  |  |  |  | **53.58% Positivity** |
| 5 MLD; 3.29 MLD | 87.35% | 14 | May 26-June 9, 2022 | 118.81% | 7 | June 16-June 23, 2022 |
| **Yelahanka Ph-I (100000)** |  |  |  |  |  | **68.97% Positivity** |
| 10 MLD; 10.21 MLD Inflow | 38.35% | 7 | May 31-June 7, 2022 |  |  |  |
| **Hulimavu (100000)** |  |  |  |  |  | **57.69% Positivity** |
| 10 MLD; 7.49 MLD Inflow | 82.17% | 14 | May 19-June 2, 2022 | 49.42% | 7 | June 16-June 23, 2022 |
| **Yele Mallappa Chetti Kere (150,000)** |  |  |  |  |  | **65.38% Positivity** |
| 15 MLD; 14.66 MLD Inflow | 50.09% | 14 | May 25-June 8, 2022 |  |  |  |
| **Jakkur (150,000)** |  |  |  |  |  | **43.33% Positivity** |
| 15 MLD; 15.34 MLD Inflow | 103.75% | 14 | May 27-June 10, 2022 |  |  |  |
| **K R Puram (200,000)♣** |  |  |  |  |  | **68% Positivity** |
| 20 MLD; 16.21 MLD Inflow | 32.20% | 14 | May 25-June 8, 2022 | 33.33% | 7 | June 15-June 22, 2022 |
| **Nagasandra (200,000)** |  |  |  |  |  | **45.83% Positivity** |
| 20 MLD; 13.48 MLD Inflow | 100.52% | 14 | May 23-June 6, 2022 | 61.65% | 7 | June 6- June 13, 2022 |
| **Nagasandra Ph-I (200,000)** |  |  |  |  |  | **36.36% Positivity** |
| 20 MLD; 10.2 MLD Inflow | 338.96% | 14 | June 13-June 27, 2022 |  |  |  |
| **Horamavu Agara (200,000)** |  |  |  |  |  | **58.33% Positivity** |
| 20 MLD; 18.69 MLD Inflow | 267.42% | 7 | May 25-June 1, 2022 | 83.57% | 7 | June 15-June 22, 2022 |
| **Agaram (350000)♣** |  |  |  |  |  | **61.54% Positivity** |
| 35 MLD; 33.8 MLD Inflow | 75.91% | 21 | May 18-June 8, 2022 | 32.83 % | 7 | June 15-June 22, 2022 |
| **Rajacanal (400000)** |  |  |  |  |  | **70% Positivity** |
| 42 MLD; 42.92 MLD Inflow | 35.53% | 7 | May 3-May 10, 2022 |  | 4 |  |
| **Doddabelee (400000)** |  |  |  |  |  | **43.48% Positivity** |
| 40 MLD; 40.46 MLD Inflow | 122.06% | 14 | May 16-May 30, 2022 |  |  |  |
| **Kadabeesanahalli Ph-I (500000)** |  |  |  |  |  | **56% Positivity** |
| 50 MLD; 42.94 MLD Inflow | 52.81% | 14 | June 16-June 30, 2022 |  |  |  |
| **KC Valley 2 (600000)♣** |  |  |  |  |  | **57.69% Positivity** |
| 60 MLD; 60.72 MLD Inflow | 26.86% | 7 | May 11-May 18, 2022 | 93.71% | 7 | June 15-June 22, 2022 |
| **Kengeri (600000)** |  |  |  |  |  | **58.33% Positivity** |
| 60 MLD; 34.29 MLD Inflow | 50.55% | 7 | May 30-June 6, 2022 |  |  |  |
| **Mailasandra Ph-I (750000)** |  |  |  |  |  | **57.69% Positivity** |
| 75 MLD; 64.9 MLD Inflow | 58.32% | 14 | May 23-June 6, 2022 |  |  |  |
| **Hebbal (1000000)** |  |  |  |  |  | **66.67% Positivity** |
| 100 MLD; 78.52 MLD Inflow | 65.69% |  | April 26-June 7, 2022 |  |  |  |
| **Bellandur Amani Kere (900000)** |  |  |  |  |  | **76% Positivity** |
| 90 MLD; 94.7 MLD Inflow | 49% | 14 | April 28-May 12, 2022 | 23.38% | 7 | May 12-May 19, 2022 |
| **K & C Valley 1 (2480000)♣** |  |  |  |  |  | **73.08% Positivity** |
| 248 MLD; 219.2 MLD Inflow | 37.10% | 7 | May 25-June 1, 2022 | 33.10% | 7 | June 15- June 22, 2022 |
| **V. Valley\* (1800000)** |  |  |  |  |  | **31.25% Positivity** |
| 180 MLD; 83 MLD Inflow | 77.56% | 21 | June 2-June 23, 2022 |  |  |  |
| **Kadugodi\* (60000)** |  |  |  |  |  | **27.27% Positivity** |
| 6 MLD; 3.57 MLD Inflow |  |  |  |  |  |  |

♣ sewershed sites showed 'red alert' with viral load crossing the upper-class limit (UCL) \* sewershed sites with PCR inhibitors/poor quality data.

The viral trend showed an increase from 15^th^ April 2022 which remained below the early warning stage, with a consistent 30-70% rise in the following weeks reaching the ‘early warning’ stage by 17^th^ June 2022 (Table 1).

We detected comparable levels and variable SARS-CoV-2 infection dynamics for STPs (Fig. 4). All STPs showed viral signals throughout the study period. However, 10 STPs consistently showed high positivity (Table 1). The EWMA varied for each STP driven by the capacity, inflow rate and catchment area. Four sewershed sites-Agaram, KR Puram, KC Valley1 and KC Valley2 showed ‘red alert’ with the EWMA viral load greater than the projected line (upper control limit). Seventeen sewershed sites; Chikkabegur, Chikkabanvara, Cubbon Park, Doddabelee, Hebbal, Horamavu Agara, Kengeri, Mallathahalli, Rajacanal, V. Valley, Sarakki, Hulimavi, Yele Mallappa Chetti Kere, Nagasandra, Nagasandra-Ph-1, Kadabeesanahalli Ph-1, and Bellandur Amani Kere showed early warning signal with viral load projected above the mean line (Fig. 4). The EWMA mirrored estimated infected individuals for the catchment area of each STP and mainly reflected with an increase in estimated infected individuals with an increasing viral trend.

**Fig. 4.**
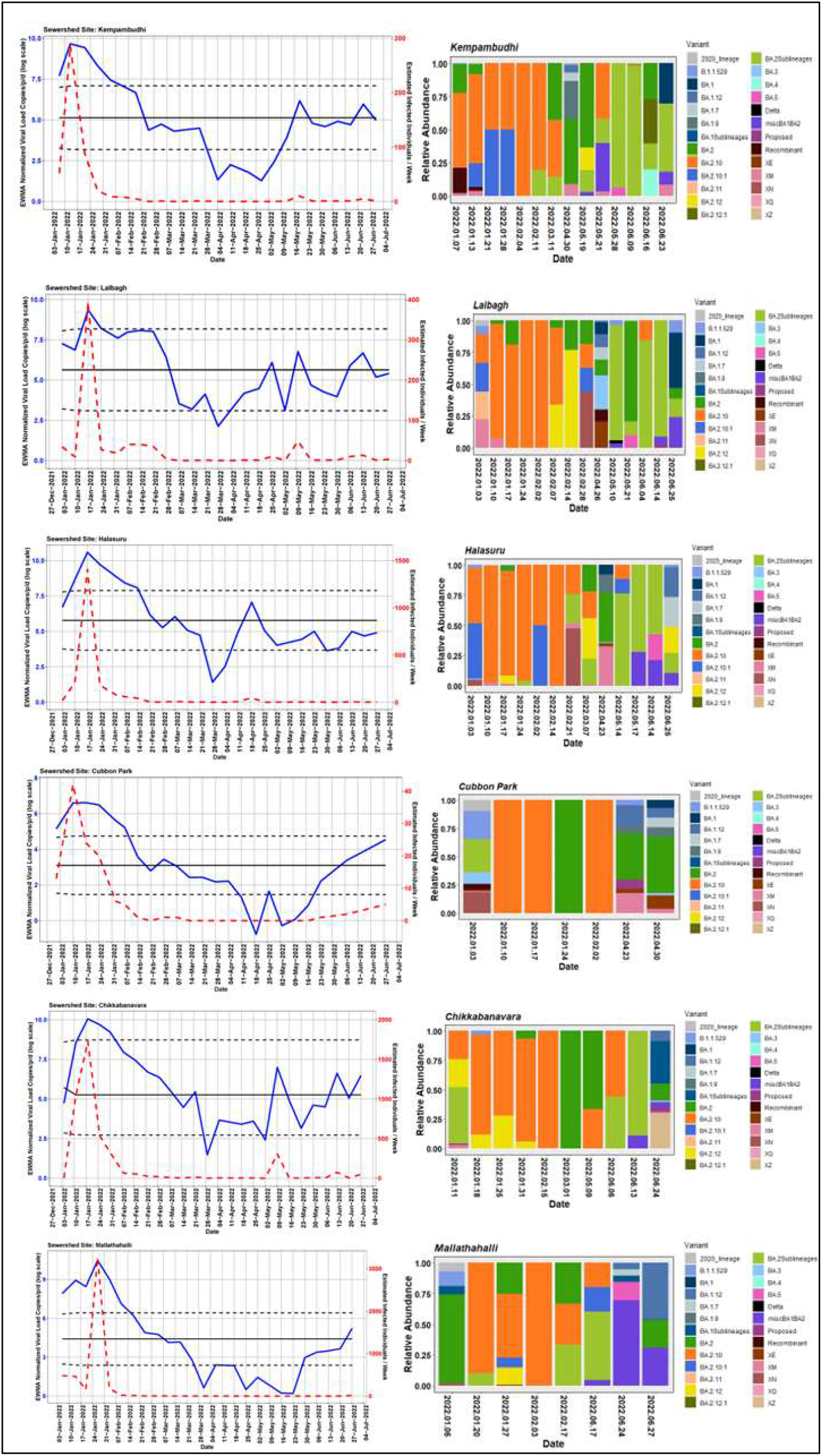
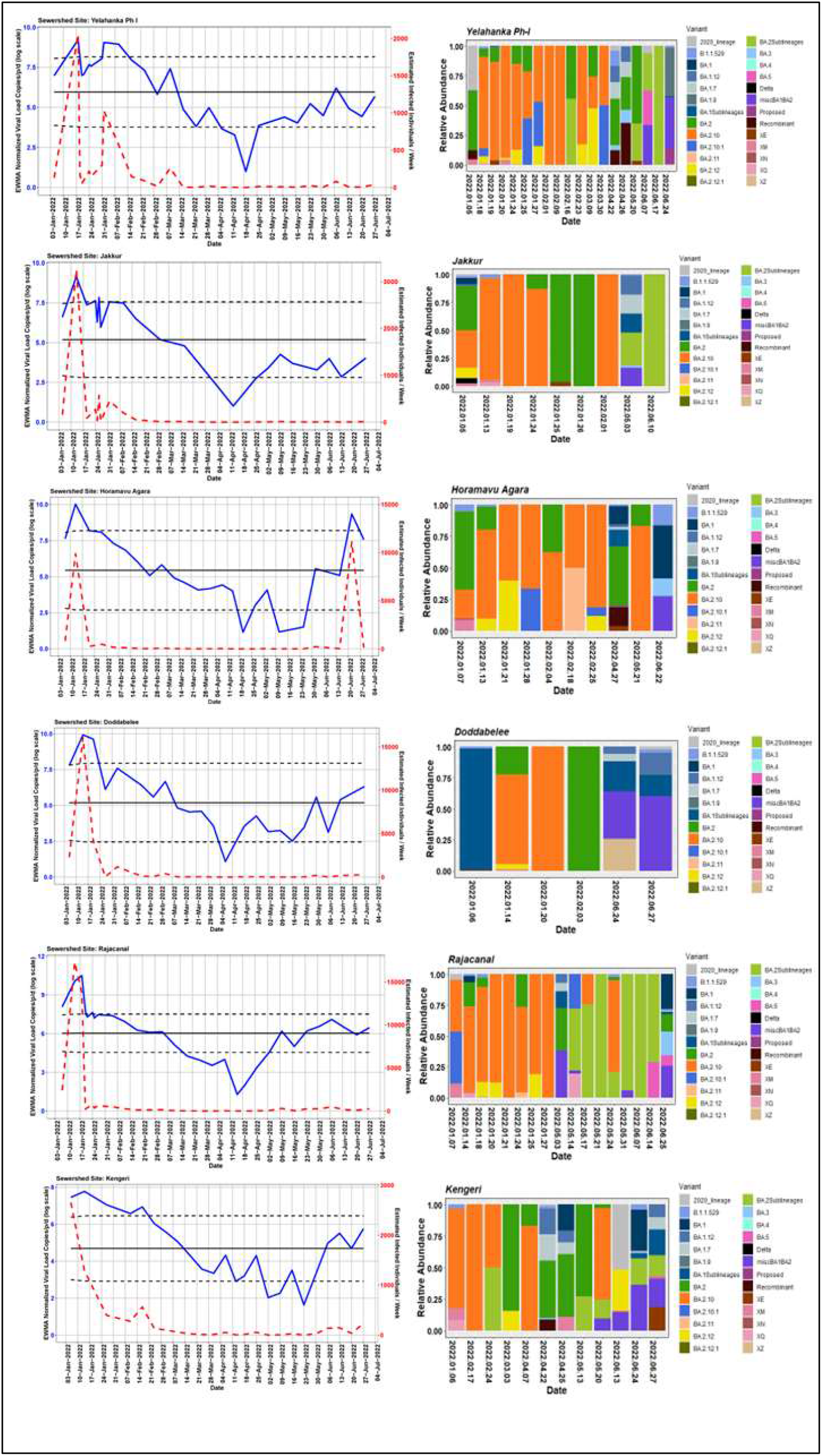
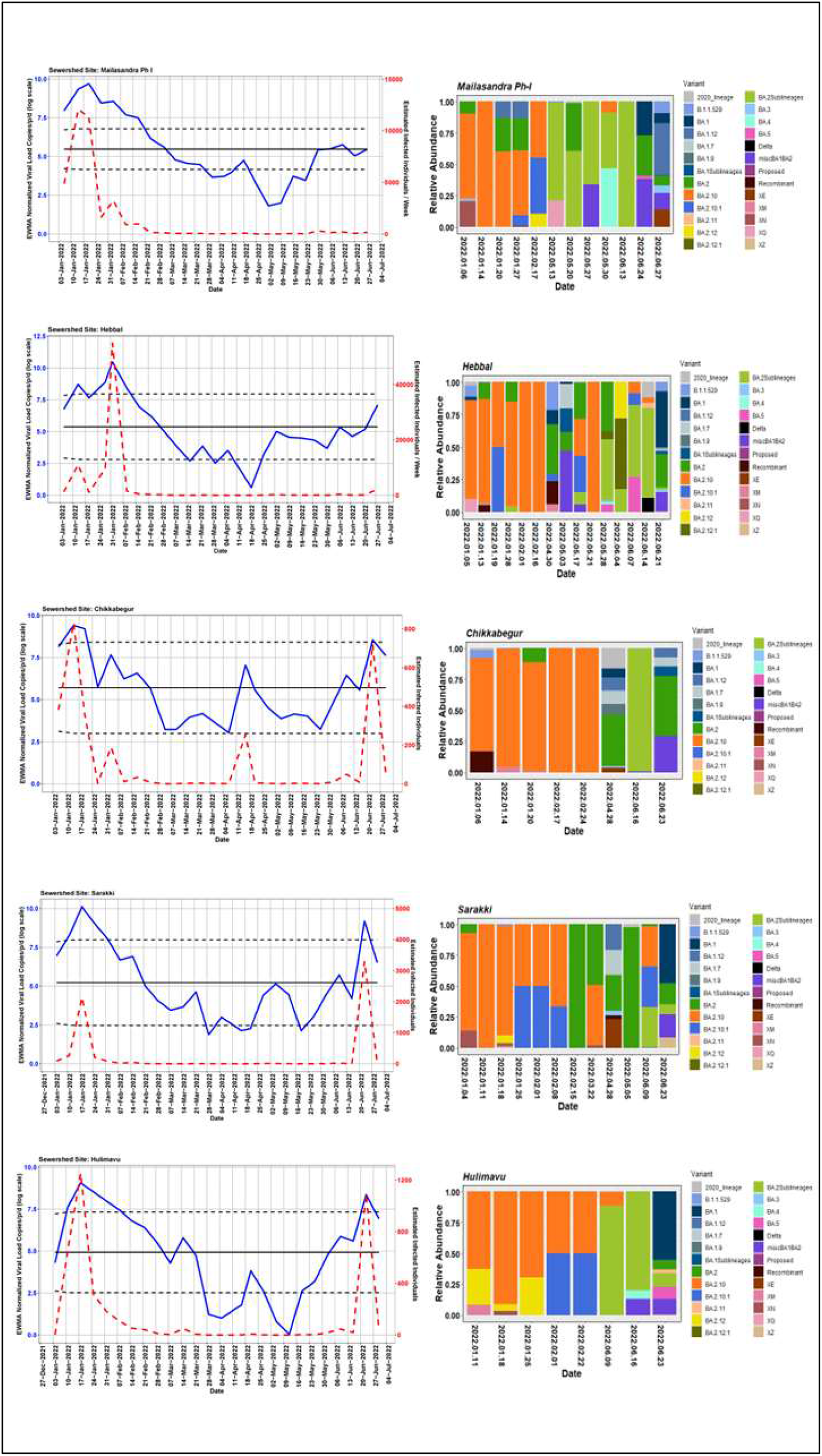
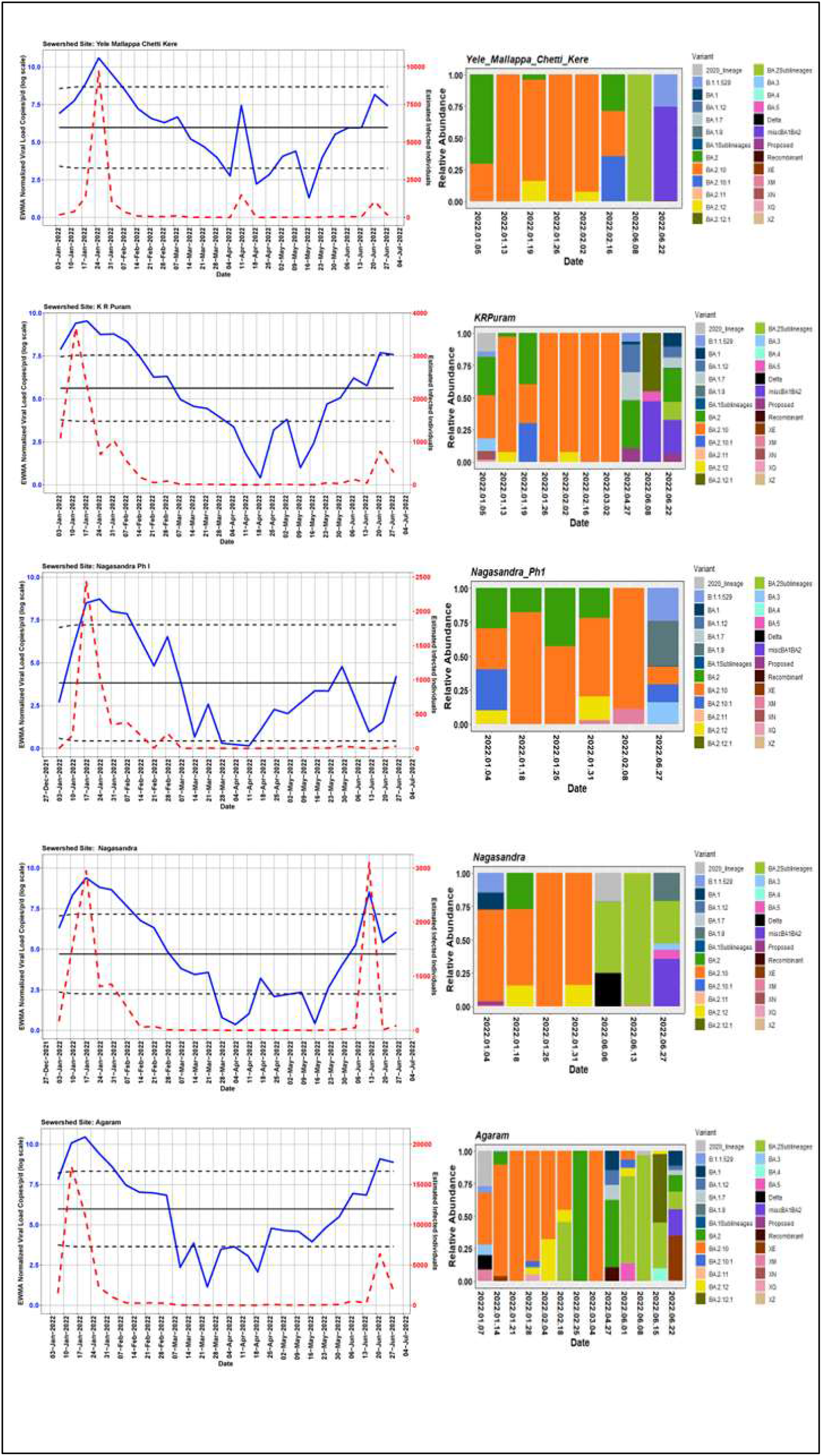
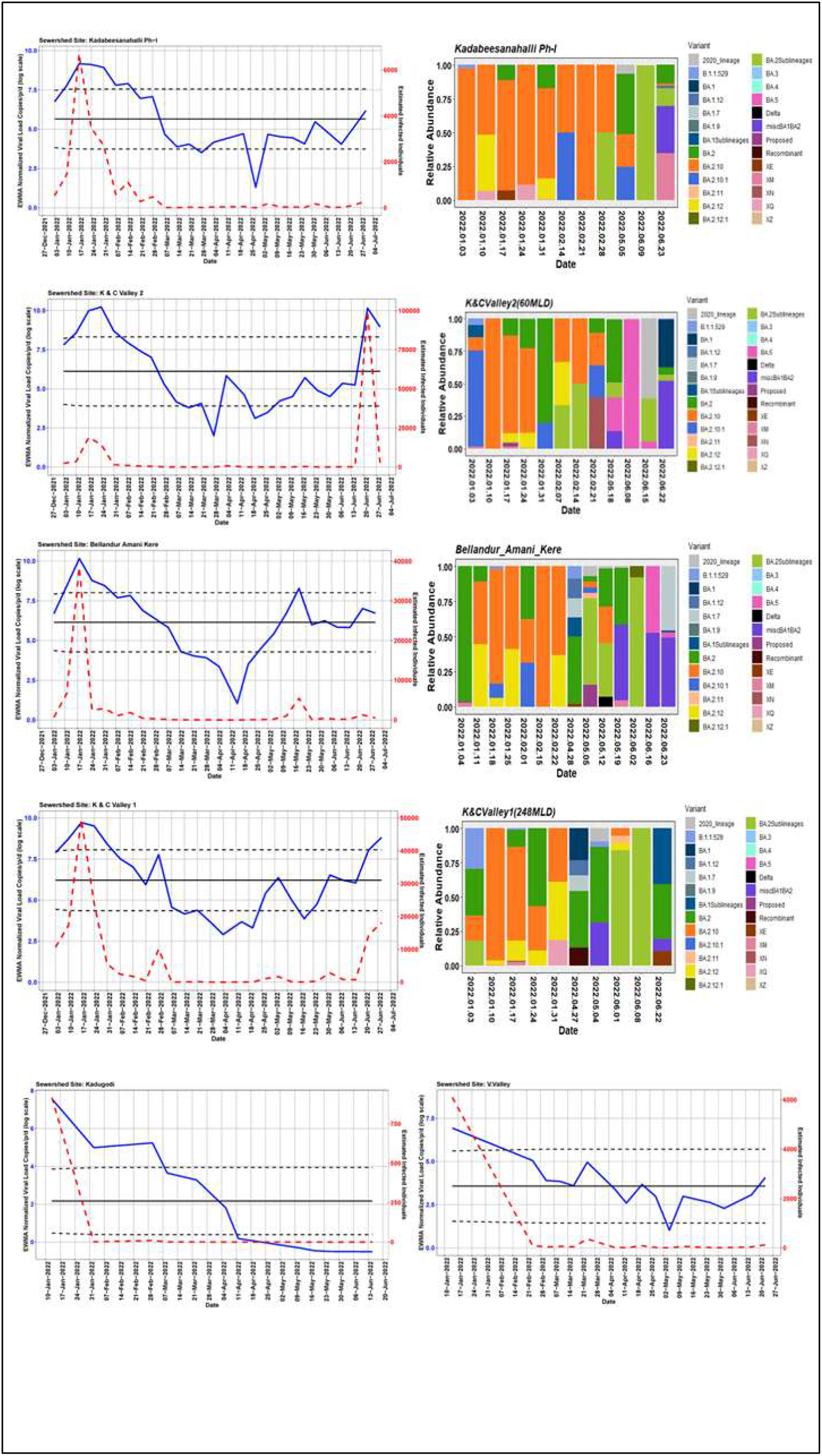
The exponentially weighted moving average charts for each sewershed site and the relative abundance of variants by date of collection.

### SARS-CoV-2 abundance and diversity in wastewater

We analysed 878 wastewater samples from 1 January 2022-30 June 2022. To compare wastewater genomic surveillance with clinical surveillance, we sequenced 422 positive wastewater samples. The genome coverage ranged between 1%–99%. The CT values showed a negative correlation with viral load copies (–51.34, p<0.0001) and genome coverage (–1.77, p<0.0001) (Fig. S1). Therefore, we considered samples (n=304) above 70% genome coverage for subsequent analyses. The low viral copies between 28^th^ March to 8^th^ April 2022 are reflected in our sequencing success; hence, there is a gap in sequencing data despite continuous sampling. The read abundance was not significantly different across month (Fig. S2).

Using *Freyja* we analysed relative abundance and diversity in SARS-CoV-2 lineages. Our validation results for clinical samples showed the expected lineages. In addition, a mixture of variants that were not identified using traditional pipeline (see Fig. S3). These clinical samples were reported infected with only one variant, however, we found mutations associated with other variant suggesting mixed infections. Our follow-up analysis using *Frejya* showed a total abundance of 1220 lineages representing 152 distinct lineages in wastewater from January-June 2022 (Fig. 5). Of these seven lineages from Omicron family were in dominant. We found BA.2.10 was the most dominant lineage (14.83%), followed by BA.2 (10.49%), B.1.1529 (5.1%), BA.2.12 (5%) and BA.2.10.1 (3.1%). We found signature mutations of two recombinant lineages of BA.1.1 and BA.2 - XM (3.9%) and XQ and XE (1%).

**Fig. 5.**
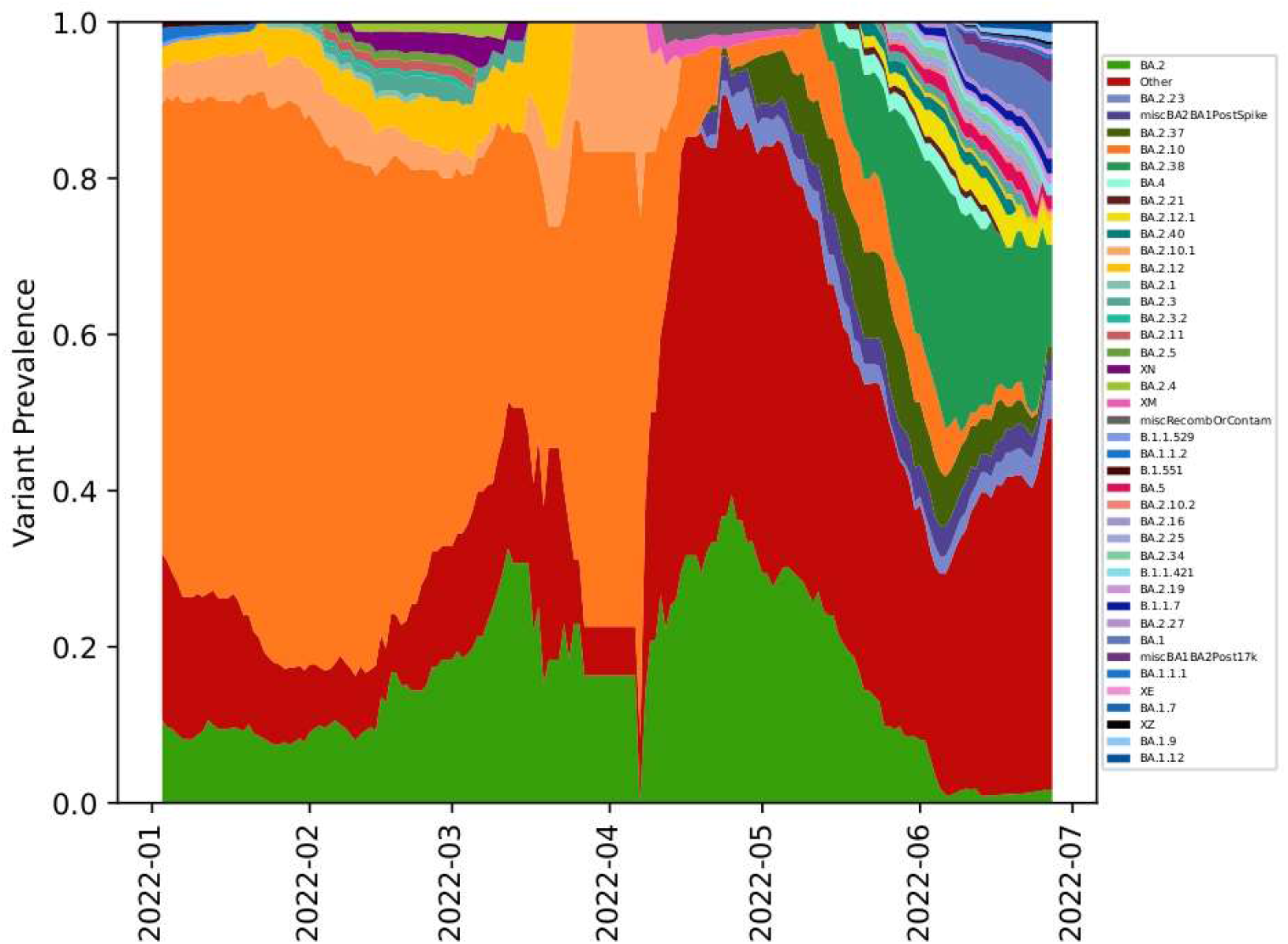
Relative abundance of SARS-CoV-2 lineages in wastewater by date of collection in Bangalore.

We analyzed the Shannon indices to understand overall alpha diversity in SARS-CoV-2 which was significantly different between months (*F*_[5, 253]_ = 19.6, *P* < 0.001) and then used Tukey’s honestly significant difference (HSD) *post hoc* pairwise comparison testing to show that only January-April, February-April, June-April, March-April, June-February, May-February, June-January and May-January alpha diversities were different from each other (adjusted *P* value [*P*_adj_] < 0.05) (Fig. 6). In contrast, the alpha diversity was not significantly different between STP (*F* _[27, 201]_ = 0.65, *P* = 0.90). We also compared the Bray-Curtis dissimilarities of the samples with Adonis and found that overall beta diversity values by month were significantly different (*R*^2^ = 0.27, *P* < 0.001) but not by STP (*R*^2^ = 0.09, *P* =0.93).

**Fig. 6.**
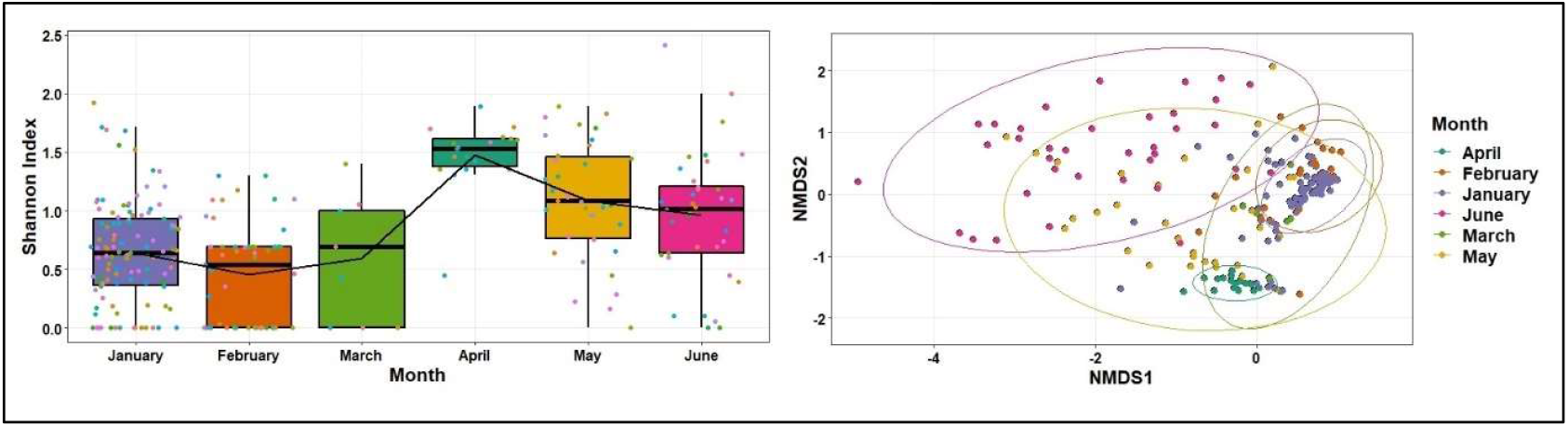
Shannon diversity (alpha diversity) in lineages across months and non-metric multidimensional scaling (NMDS) ordination of the Bray-curtis dissimilarities in lineages by month.

Finally, we used the linear-mixed effect model to show that the relative abundance of seven dominant lineages (VOC) showed change over time - BA.2, XM (recombinant lineage of BA.1.1 and BA.2) and XQ showed an increasing trend whereas BA.2.10, BA.2.10.1 decreased from January to June. BA.2 lineage remained dominant in May along with ‘other’ lineages found in low proportions (<1%). B.1.1.529 and BA.2.12 showed no significant change in their relative abundance (Fig. 7). BA.2.12 was the dominant lineage during the third wave in January with high viral load copies/ml.

**Fig. 7.**
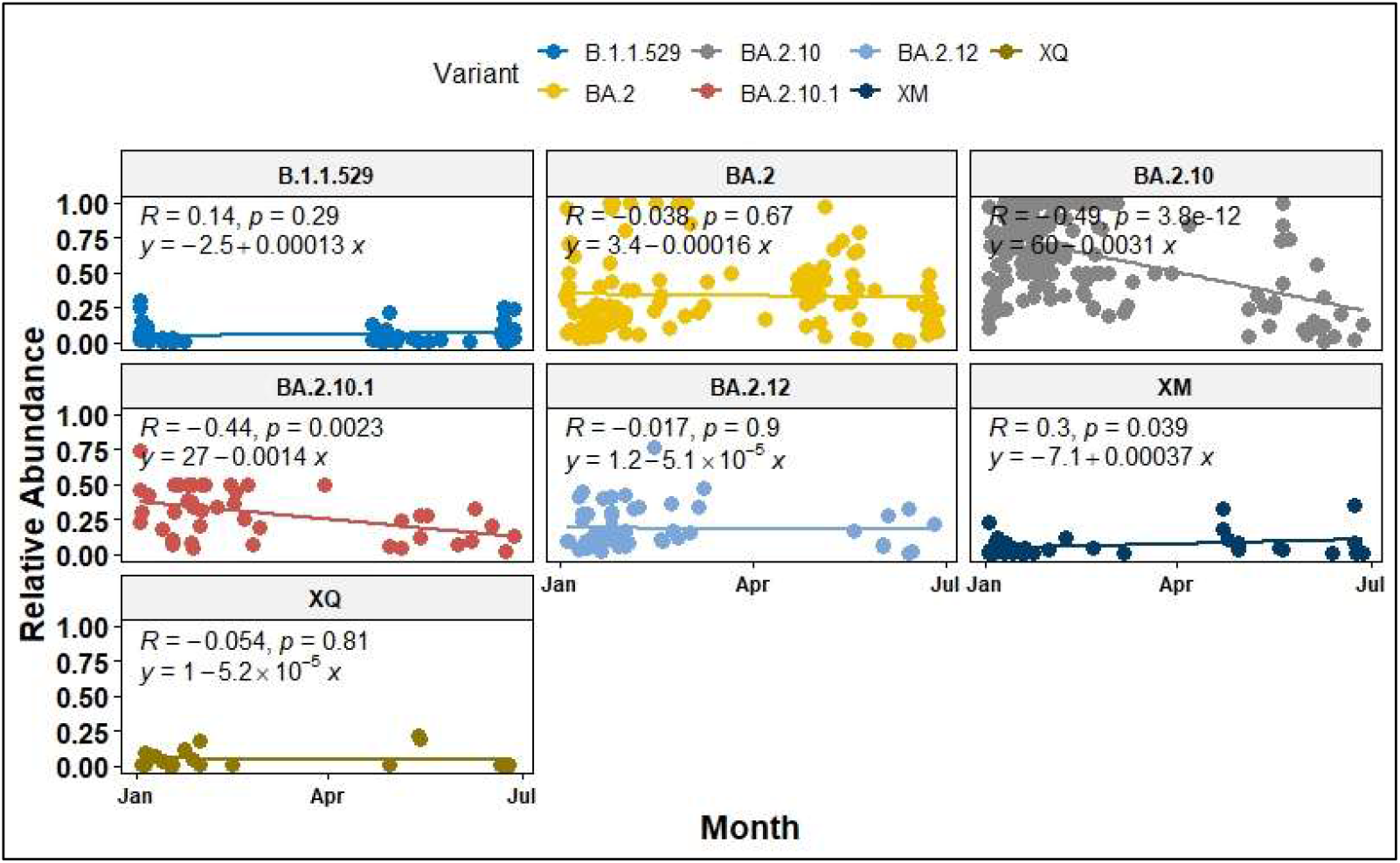
Relative abundance of dominant SARS-CoV-2 lineages showed a variable trend between January to June 2022.

### Comparison between Wastewater and clinical data

We analyzed 13,478 SARS-CoV-2 genomes submitted on GISAID from January to June (January; 7827, February; 3364, March; 364, April; 229, May; 642, June; 1052) from Bangalore. Of these, 71 were distinct lineages that followed a similar pattern as wastewater samples-BA.2.10 was the dominant lineage with 51.65% of the genomes, 26.90% of the genomes were assigned to BA.2 and ‘unassigned’ (recombinant lineages) constituted 8.73 % of the diversity.

To test whether wastewater genomic surveillance can detect changes in lineage abundance circulating at the community level, we compared dominant Variant of Concern (VOC; variants associated with high transmissibility or immune evasion) detection rates between clinical and wastewater sequenced data. We found that the general trend in lineage abundance remained very comparable to the dynamics between wastewater and clinical data. Furthermore, the wastewater genomic surveillance consistently recorded mutations associated with ‘other’ lineages in low frequency (<1%), which were otherwise not seen in the clinical samples. There was a huge mismatch in lineage diversity which was significantly higher in wastewater than recorded in clinical samples (Mann-Whitney U test, p<0.001; Fig. S4).

To understand if wastewater sampling can help in the early detection of emerging variants in the city, we compared the first detection of VOC in wastewater with the collection dates of clinical samples sequenced as part of genomic surveillance in Bangalore. In contrast, the exact collection dates for many samples are not mentioned in the database, which prevents a temporal comparison across several lineages. Nonetheless, we compared the month of the emergence of VOC in clinical samples from GISAID. For example, BA.2.10.1, BA.2.12 were detected two months prior in wastewater in January 2022 to the first detection in clinical samples in March 2022 (Fig. 8).

**Fig. 8.**
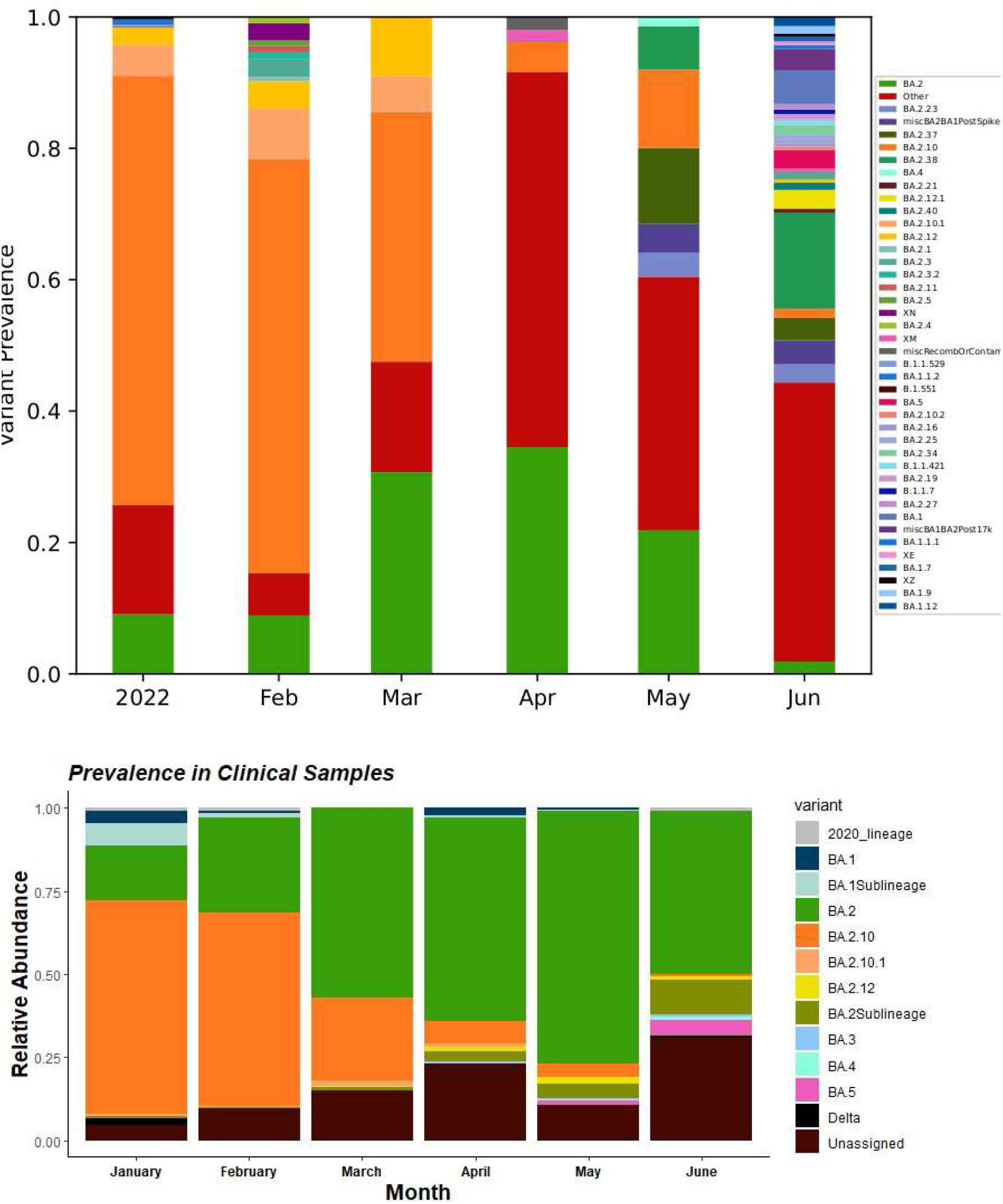
Aggregated relative abundance of SARS-CoV-2 variants by month in wastewater in the upper panel analysed using *Freyja* and in clinical genomic surveillance in the lower panel. Lineages retrieved in less than 1% relative abundance are aggregated as ‘Other’. **BA.1Sublineages** are aggregate of lineages less than 1% – BA.1.1, BA.1.1.1, BA.1.1.16, BA.1.1.7, BA.1.14, BA.1.15, BA.1.15.1, BA.1.17, BA.1.17.2, BA.1.18, BA.1.20. **BA.2Sublineages** are aggregate of lineages less than 1% – BA.12.1, BA.2.18, BA.2.20, BA.2.21, BA.2.23, BA.2.27, BA.2.3, BA.2.3.1, BA.2.31, BA.2.32, BA.2.38, BA.2.4, BA.2.40.1, BA.2.9. See Fig. S4 for details.

Until June end, there were 12 genomes of BA.4, and 58 genomes of BA.5 were submitted on GISAID. The first clinical sample of BA.5 was collected on 11 May 2022 and sequenced in wastewater sample collected on 18^th^ May 2022 from two STPs (Lalbagh and KC Valley2). BA.5 was detected on two consecutive sampling on 8^th^ June and 15^th^ June from KC Valley2. Similarly, BA.4. was first isolated on 24^th^ May 2022 in clinical samples and first found in wastewater on 28^th^ May 2022 from Hebbal sewershed site and 30^th^ May 2022 from Mailasandra subsequently sequenced in a sample collected on 16^th^ June in Hulimavu. In total, 73 spike mutations were recorded in BA.4 and BA.5 in the wastewater sample (Fig. S5). The increases in lineage detection frequency for VOC - ‘unassigned’ (recombinant lineages probably equates to XM, XE, XN in wastewater) showed an increasing trend whereas BA.2.10 and BA.2 declined with time.

Our study shows a localized increase in variant diversity and a spike in VOC abundance -BA.1, BA.1 sub lineages, BA.2 sub lineages, and prevalence of ancient SARS-CoV-2 lineages recorded in 2020 and Delta lineages across STPs. In addition, the lineage richness was higher in the ‘red alert’ stage of an emerging wave than during the ‘wave’ phase (see Fig. 2). Both wastewater and clinical datasets showed low viral richness in March-April. However, there was a huge surge in lineage diversity in June which corresponded with increasing viral load both at citywide as well as STPs. This further highlight that the increasing trend was contributed by a shift from BA.2.10 to many BA.2 sub-lineages rather than one dominating lineage at the community level. BA.2.10 was the most dominant lineage seen in both wastewater and clinical samples during the peak of the third wave in January 2022 (Fig. 5).

## Discussion

Our longitudinal study provides an in-depth analysis of the SARS-CoV-2 viral concentration and how this relates to the lineage dynamics in wastewater capturing data from more than 11 million people in Bangalore city. The strong correlation between wastewater viral concentrations and daily reports of new clinically confirmed COVID-19 cases (Fig. 3) – combined with the time lag between the wastewater signal and clinical data – suggests that newly infected individuals contribute significant viral loads to the wastewater. This further suggests that that most of this shedding may occur early in infection (asymptomatic phase), prior to the individual seeking healthcare and being tested^16^. Therefore, wastewater surveillance can be used as a complementary tool to clinical testing to predict trends in new COVID-19 cases. SARS-CoV-2 concentrations in wastewater began to increase exponentially in late March which led to a mask mandate by the 26^th^ April-this period coincided with the opening of schools and an increase in public movement with a reduced remote working. Wastewater surveillance revealed the largest number of positive cases likely to be Mahadevapura zone, East Zone, and Bommanahalli –by <u>7^th^ June</u>-there was renewed mask mandate-which corresponded with a surge in viral load in four sewersheds. We note that inadequate testing, changing criteria of testing and population seeking home tests over RTPCR could introduce uncertainty to the reported cases. Nonetheless, the time-lag used for modelling fits the data and signifies the timing of infection-that wastewater signals may reflect shedding dynamics early in infection.

Each STP showed a variable viral dynamic which could be primarily driven by individual shedding rates, viral stability in wastewater, and flow rates of the influent. Our sensitivity analysis indicated that the estimated number of infected individuals at a citywide level was four folds higher than reported cases. This information has been crucial in enhancing testing in targeted locations for the early detection of asymptomatic cases. One of the limitations of our analysis is the lack of clinical testing data from catchment areas of each sewershed site which could help correlate with viral load to devise an effective and economical strategy to track the timing, location, and magnitude of SARS-CoV-2 activity outbreaks. Two sewershed sites-Kadugodi and V. Valley had technical challenge in obtaining high quality data and sequences from samples with low viral load and elevated levels of PCR inhibitors.

Wastewater data is often fragmented and does not capture all variant-defining mutations on a single genome. Nonetheless, with an increase in detection of multiple variant-associated mutations or mutations not shared by other known variants, we classify these lineages as variant-like upon following *Frejya* which used the UShER tree with WHO designation and *outbreak*.*info* metadata^37^. Our wastewater samples contained a diversity of SARS-CoV-2 lineages that showed significant difference across months but not by sewershed sites suggesting that there was no geographical variation in viral composition in the city. Furthermore, we found low proportion of lineages which were not reported in the clinical data-this discrepancy highlights that clinical testing and sequencing as per the national guidelines, only symptomatic individuals are tested. There were lineages only found in clinical samples but not seen in the wastewater which probably suggests that a very low number of individuals harboured those variants. This further signifies that dominant VOC infecting a large proportion of individuals exhibited similar trends in the wastewater. Furthermore, the clinical data is analysed using a traditional pipeline which assigns a single variant based on the dominant mutations in the sample. We used a pipeline designed to calculate proportions of VOC from the environmental samples that is a highly reproducible computational analysis pipeline with comprehensive reports. Our validation of the pipeline using clinical samples showed mutations associated with mixed lineages in clinical samples (Fig. S3) which probably highlights that the diversity in clinical data might be underestimated and the need to consider alternate approaches.

Given that we sequenced only 2% of wastewater samples relative to the sequenced clinical samples, the emergence of variants in wastewater followed a trend that matched with other countries. For example, the rise of BA.5 is occurring at the same time as the decline in BA.2 in several countries. We detected BA.2.10.1, BA.2.12 two months prior in wastewater in January 2022 to the first detection in clinical samples in March 2022 in Bangalore whereas BA.4 and BA.5 were detected in 4-7 days late in wastewater. The emergence of a variant in the wastewater implies that a significant proportion of individuals in the community are infected with that variant and shedding the virus. Wastewater testing can provide a less biased snapshot of viral diversity and community health^17^. Whereas a late detection in the clinical sample could happen due to limited or biased testing, sequencing^18^ or a large proportion of individuals were asymptomatic or home testing upon COVID-19 symptoms. Clinical samples were sequenced from a selected hospital which is not representative of the Bangalore population. Karthikeyan et al.^19^ observed varying periods of VOC lineage detection relative to clinical genomic surveillance, which was attributed to different virus shedding characteristics across lineages^20^.

While we saw a temporal trend in variants, there was no geographical structure in lineages across sewershed sites. Nonetheless, we noticed a similar pattern in viral load and shift from BA.2.10 to BA.2 sub-lineages-we observed a strong positive association between the local VOC frequency and estimated number of infected individuals which were consistent with the increased transmissibility of this VOC identified.

Our study highlights that quantifying viral titres, correlating with the known number of cases in the area combined with genomic surveillance helps in tracking of VOCs over time and space and helps inform policy-making decisions in order to control new outbreaks. Several WBE initiatives for SARS-CoV-2 monitoring were established worldwide, and currently, the COVIDpoops19 initiative^1^ lists 128 dashboards and our data is displayed as part of the Bangalore pandemic response initiative. The findings from this study have been discussed regularly with Bruhat Bangalore Mahanagara Palike (BBMP) and Bangalore Water Supply and Sewerage Board (BWSSB) to inform policy-making decisions. Our approach can support establishing WBE for monitoring and early-warning system for detecting pathogens beyond SARS-CoV-2.

## Conclusion

We developed an early warning system, and its performance was demonstrated by detecting and tracking the SARS-COV-2 infections and variant diversity in the local community and city levels from January 2022 to June 2022. The EWMA chart is able to capture the historical COVID-19 infection patterns and distinguish between endemic situations and the outbreak patterns. Real-time genomic surveillance is the key to understanding the emerging patterns in viral load and variants in the city as it helps to develop a pandemic plan as well as be prepared for future pandemics.

## Materials and Methods

### Wastewater sample collection

We sampled influent wastewater from 28 sewage treatment plants (STPs) in Bangalore. Samples were collected once a week from each STP (January-June 2022) and twice a week from 14 STPs (April-June 2022). For a constant comparison across the sites, we consider only once a week sampling period in this study. The details of inflow rate, STP capacity (volume of water), population size is provided in Table 1 and Figure 1.

Samples were collected in 200ml plastic bottles, tightly sealed upon collection, and stored at 4°C in the field. All samples were processed within 24 hrs at a biosafety level 2 facility following protocol (modified from^11-13, 20-23^) to provide near real-time information on viral concentrations in sewage. Briefly, samples were subjected to heat inactivation and incubated in a 200 ml bottle at 60 degrees for 90 minutes and divided into three replicates. Subsequently, the supernatant from the master samples (200 ml bottle) was transferred to the 50 ml centrifuge tube containing 0.9 gm NaCl and 4 gm PEG (8000 MW). The PEG and NaCl mixture in the samples was vortex until it dissolved, and the solution was centrifuged at 11000 rpm for 30 mins. After discarding the supernatant, 600 µl of lysis buffer was added to resuspend the pellets. Finally, the solution was transferred to a centrifuge tube, briefly vortexed to dissolve the pellet. Viral RNA was extracted following Qiagen Viral RNA mini kit protocol and 50ul of elution was stored at -80°C until subsequent analysis.

### Viral Reverse Transcriptase-quantitative PCR (RT-qPCR)

Using GenepPath Dx CoViDx One v2.1.1TK-Quantitative multiplex RT-qPCR kit, each sample was screened for the presence of SARS-CoV-2 RNA. The kit targets three viral genes: N-gene, RdRp-gene, and E-gene along with a human control gene (RNAase P gene). Since the kit is designed for a qualitative result of positive or negative, for the quantitation of viral load in samples, the <u>Covid-19 Viral Load Calculation Tool</u> (RUO) was used. A negative extraction control and extraction control was analyzed with each plate. In order to eliminate the false negative, RT-qPCR was performed on three extracted replicates of each sample and any replicate that generated a result defined as ‘positive’ by the test manufacturer was considered positive. Samples with invalid results were repeated as per the manufacturer’s instructions.

### Early warning SARS-CoV-2 detection modelling

By investigating viral RNA concentrations in the wastewater and correlating it with community testing data on a weekly basis, we generated an early and real-time map of COVID-19 infection dynamics in the city. We conducted two main analyses using the raw and normalised viral load. At the citywide level, we calculated the daily viral load, *VL* (copies/d) for SARS-CoV-2, by normalizing *C* (copies/mL) to the average daily STP flow, *Q(L/d)* using equation 1^23^) and P is the population of inhabitants the catchment of the respective sewershed.

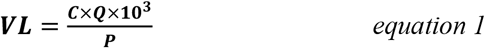

### i) Spatial and temporal dynamics in viral load copies by STPs and at the citywide level

To understand spatial and temporal changes in viral load, we used an Exponentially Weighted Moving Average (EWMA)^23^ as the monitoring algorithm to detect moderate and persistent shifts in cases. The EWMA is calculated as follow:

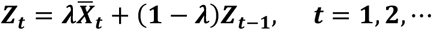

Where, *λ* is the smoothing parameter with condition 0 < *λ* ≤ 1 and 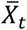 is the mean of the process at time *t*.

We used the control limits of the EWMA chart for detecting a mean shift as follow:

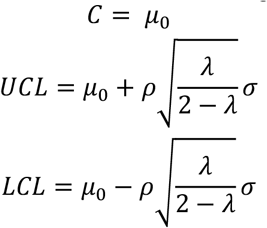

Where, *C* is the mean, *UCL* is the upper control limit, *LCL* is the lower control limit, *σ* is the process standard deviation, *ρ* is the width of the boundary of the control and EWMA chart values are computed by using the R package *“qcc*.*”*^25^

The EWMA control chart as the surveillance algorithm triggers an alarm of outbreak once the monitoring statistic exceeds the control limit, which is computed based on the design of the control chart. The EWMA gives the maximum weight to the most recent observations and exponentially gives less weight to all earlier observations. Since no previous wastewater data was available from the city, we define the threshold as the sum of the mean weekly viral load. We use the mean line as the threshold, and the ‘early warning’ signal is when the weekly viral load/incidence exceeds it. If predicted data points are below/above the average line, this indicates a less/high risk of covid -19 infection. However, if predicted data points are greater than the projected line (upper control limit), this indicates a ‘red alert’ of infection with COVID-19.

SARS-CoV-2 viral load copies in the wastewater can potentially be affected by the incubation period of a particular variant, population susceptibility, and differences in degradation rates in the sewer system. The mean incubation period of the ancestral SARS-CoV-2 variant is 6.4 days^26^, the SARS-CoV-2 Delta variant is 4.8 days^27-28^, and the Omicron variant is 3.6 days^28^. To compare the SARS-CoV-2 wastewater signal with COVID-19 cases at the citywide level, we used time lags of 7-days and 4-days to accommodate variable incubation periods of multiple variants seen in the wastewater (see section on variant analysis). Daily reports detailing the number of samples tested and positive for COVID-19 were collected from the BBMP COVID-19 portal (https://apps.bbmpgov.in/Covid19/en/mediabulletin.php). The omicron and delta SARS-CoV-2 variants shed culturable virus more than five days after symptom onset or first positive test^29^, and virus in feces can remain for more than 20 days, in which case prolonged shedding may contribute significant signal to wastewater^29-30^. We modelled EWMA with time lags of 4-days and 7-days. Time-step (4-days and 7-day lag) Pearson’s R correlation analyses were performed to evaluate the fit between log-transformed SARS-CoV-2 viral copies in wastewater and reported clinical data. We did not conduct this analysis at the STP level as the clinical testing data for each sewershed catchment size is unavailable. Using citywide clinical testing data gives a biased estimate due to varying inflow and catchment size of each sewershed.

#### ii) Estimation of infected individuals by STP and citywide

We estimated the number of infected individuals by each STP and citywide. The prevalence of SARS-CoV-2 infection within the catchment of each STP was estimated using the total number of viral RNA copies in wastewater each day, as measured in wastewater by RT-qPCR, and the number of SARS-CoV-2 RNA copies shed in stool by an infected following Ahmed et al.^10^ equation 2

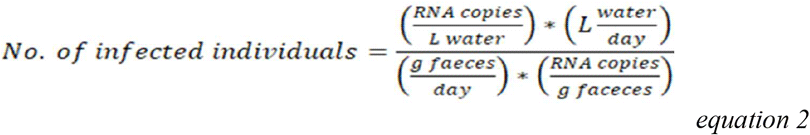

Briefly, SARS-CoV-2 RNA copies/L of wastewater were modelled as point estimates for each date of detection. The daily flow rate of wastewater was calculated for each sewershed using the product of the population of each catchment area and the observed average per capita wastewater rate of 100L/person/day. The daily stool mass of 128 g was used as per^31^ and one person sheds SARS-CoV-2 RNA 10^7^copies/g of feces as per^32^. Spearman’s correlation coefficient was used to calculate the sensitivity of the estimated number of cases and normalized viral load of each sewershed.

### SARS-CoV-2 genome amplification and sequencing

All samples positive with the RTqPCR kit were sequenced at the Next Generation Genomics Facility in National Centre for Biological Sciences, Tata Institute for Fundamental Research, Bangalore. The libraries were prepared using the Illumina COVIDSeq Test kit (Cat no: 20043675, Illumina Inc, USA). Extracted RNA samples were primed by random hexamers for reverse transcription. The cDNA products were amplified using primers targeting the entire SARS-CoV-2 genome and human cDNA targets in two different multiplex polymerase chain reaction (PCR) reactions. The amplified product was later processed for tagmentation, and adapter ligation using IDT for Illumina Nextera DNA Unique Dual Indexes Sets A–D IDT for Illumina-PCR Indexes Sets 1–4 (384 Indexes, Cat no: 20043137, Illumina Inc, USA). Further enrichment and clean-up were performed as per the manufacturer’s instructions.

Pooled libraries were quantified using a qubit 4.0 fluorometer (Invitrogen, USA), and library sizes were analysed using TapeStation 4200 (Agilent, USA). The libraries were normalized to 2nM and denatured with 0.1N NaOH. 8.1pM of denatured libraries were loaded onto HiSeq Rapid SR flow cell v2 followed by dual indexed Hiseq 2500 - 1×50 or custom 1×120 cycle workflow as per the manufacturer’s instructions (Illumina Inc).

### Bioinformatics

Wastewater samples consist of a mixture of variants circulating in a population in contrast with clinical samples which might be infected with a single variant. In the first step, the raw reads were aligned the reference genome of SARS-CoV-2 (MN908947). The processed reads are aligned with the reference genome by BWA Mem^33^ and various coverage statistics are taken by SAMtools coverage/bedcov^34^. The alignment was used for a single nucleotide variant (SNV) calling iVar^35^. The iVar tool was used to trim the primers and generate a table for each sample with mutation frequency data and an adj p-value (Fisher’s test) for altered positions of SARS-CoV-2 from the BAM files. iVar was run with a minimum base quality filter of 15 using the reference genome of SARS-CoV-2 and the feature file Sars_cov_2.ASM985889v3.101.gff3 from NCBI. For predicting the lineage abundances, a deconvolution matrix was generated using *Freyja* (https://github.com/andersen-lab/Freyja)^18^ - a dedicated bioinformatic pipeline for wastewater analysis. From measurements of SNV frequency and sequencing depth at each position in the genome, *Freyja* returns an estimate of the true lineage abundances in the sample. We cross-validated the efficiency of the deconvolution tool in reliably inferring the mixed samples with clinical data. We first applied the pipeline to clinical data from Bhoyar *et al*.^36^ and Bangalore (derived sequences submitted to the GISAID EpiCoV database (hereafter, GISAID; https://www.gisaid.org/). *Frejya* used the UShER tree with WHO designation and *outbreak*.*info* metadata^37^.

### Virus richness and diversity

For community diversity analyses, we used lineage abundances (normalized by the read depth within-sample relative abundances using *Freyja*) to generate Shannon diversity indices and Bray-Curtis dissimilarity matrices with *vegan* v2.5-7^38^. We also ran Adonis permutational multivariate analysis of variance (PERMANOVA) tests on the distance matrices and performed nonmetric multidimensional scaling on the data and compared the relative abundances of lineages over time with “lmerTest” v3.1-3 using STP as a random effect^39^.

## Data Availability

All raw wastewater sequencing data will be available via the NCBI Sequence Read Archive under the BioProject ID PRJNXXX. Consensus sequences from clinical surveillance
are all available on GISAID.

## Code availability

Freyja is hosted publicly on github (https://github.com/andersen-lab/Freyja) and is available under a BSD-2-Clause License (doi: 10.5281/zenodo.6585067, version 1.3.7). Freyja is accessible as a package via bioconda (https://bioconda.github.io/recipes/freyja/README.html)

## Data Availability

All raw wastewater sequencing data will be available via the NCBI Sequence Read Archive under the BioProject ID PRJNXXX. Consensus sequences from clinical surveillance are all available on GISAID.

## Acknowledgements

This work has been supported by funding from the Rockefeller Foundation grant to National Centre for Biological Sciences (TIFR) and the Indian Council of Medical Research grant to (FI) Tata Institute for Genetics and Society and Tata Trusts. We sincerely thank Bangalore Water Supply and Sewerage Board for providing access to the sewershed sites and Mr. BC Gangadhar for support throughout this study. We thank Bruhat Bengaluru Mahanagara Palike, Dr. Thrilok Chandra, Special Health Commissioner, for their support and for using the findings of this study for informed decisions. Finally, we thank Pradeep N, Vikas V and Manoj S for their help with sample collections. Finally, we thank Dr. Giridhar Babu for constructive feedback and constant support throughout this study.

## Author Contributions

FI, UR and VS: Designed the study; SL: conducted EWMA modelling, SG, AN, SJ: processed the samples, conducted RNA extractions and RT-qPCR; DS: library preparation and sequencing; ND and KP: Conducted bioinformatic analysis. RM, FI secured funding. FI analyzed the data and wrote the manuscript. All authors approve the manuscript.

## Supplementary files

**Fig. S1.**
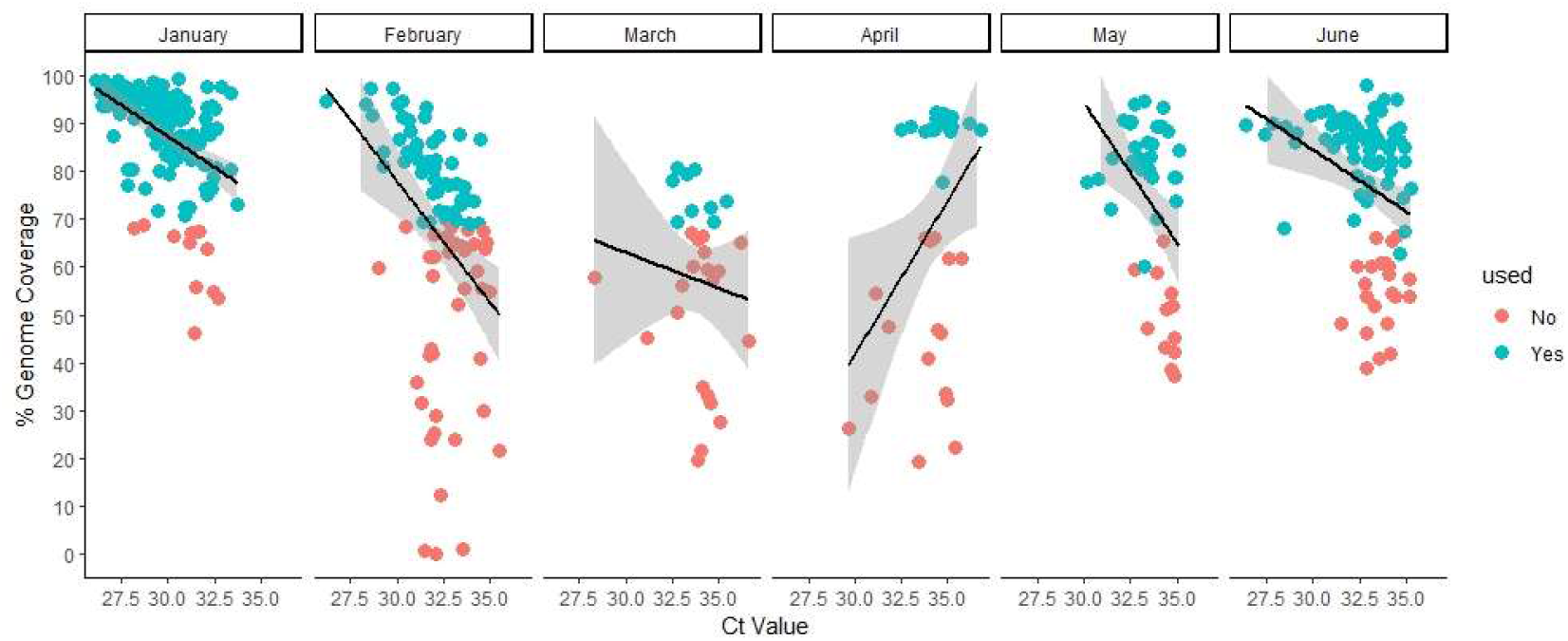
Quantitative reverse transcription PCR Ct of SARS-CoV-2 RNA in sewage samples as determined by three viral genes assays against the percentage of genome covered.

**Figure S2.**
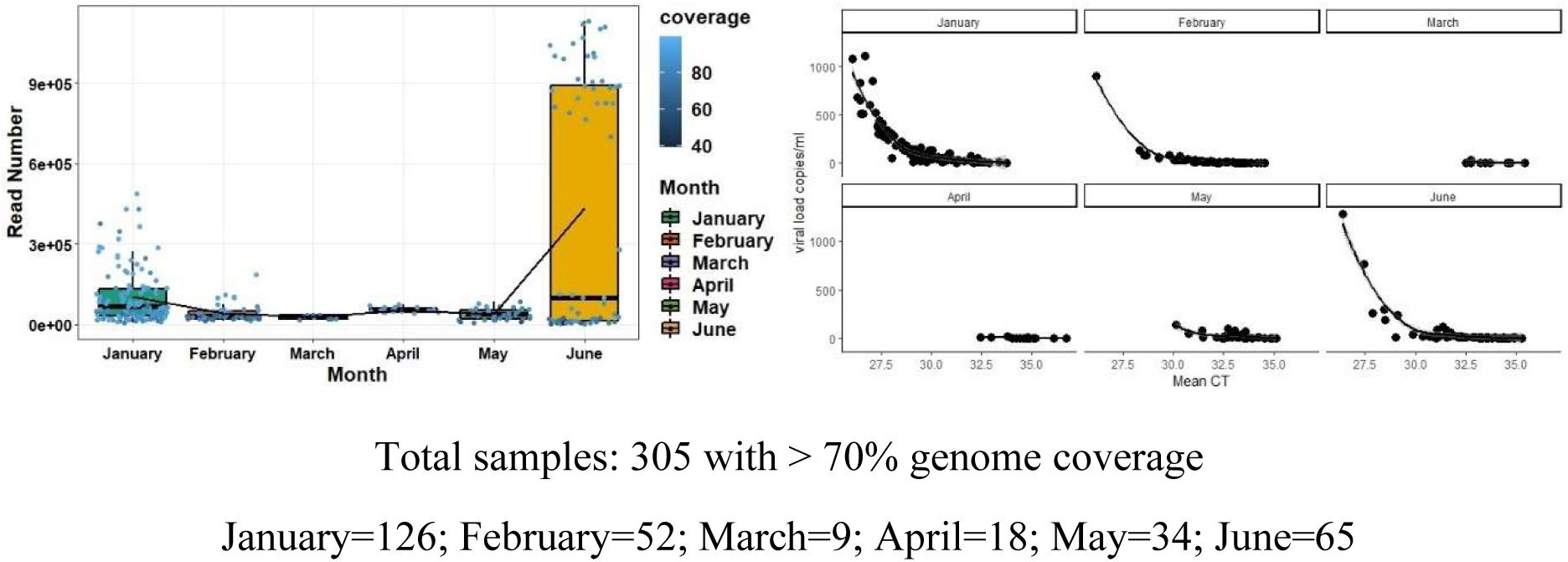
Summary of SARS-CoV-2 read abundance through time (a) and negative association between mean CT values of three viral genes and viral load copies in wastewater samples from Bangalore city.

**Figure S3.**
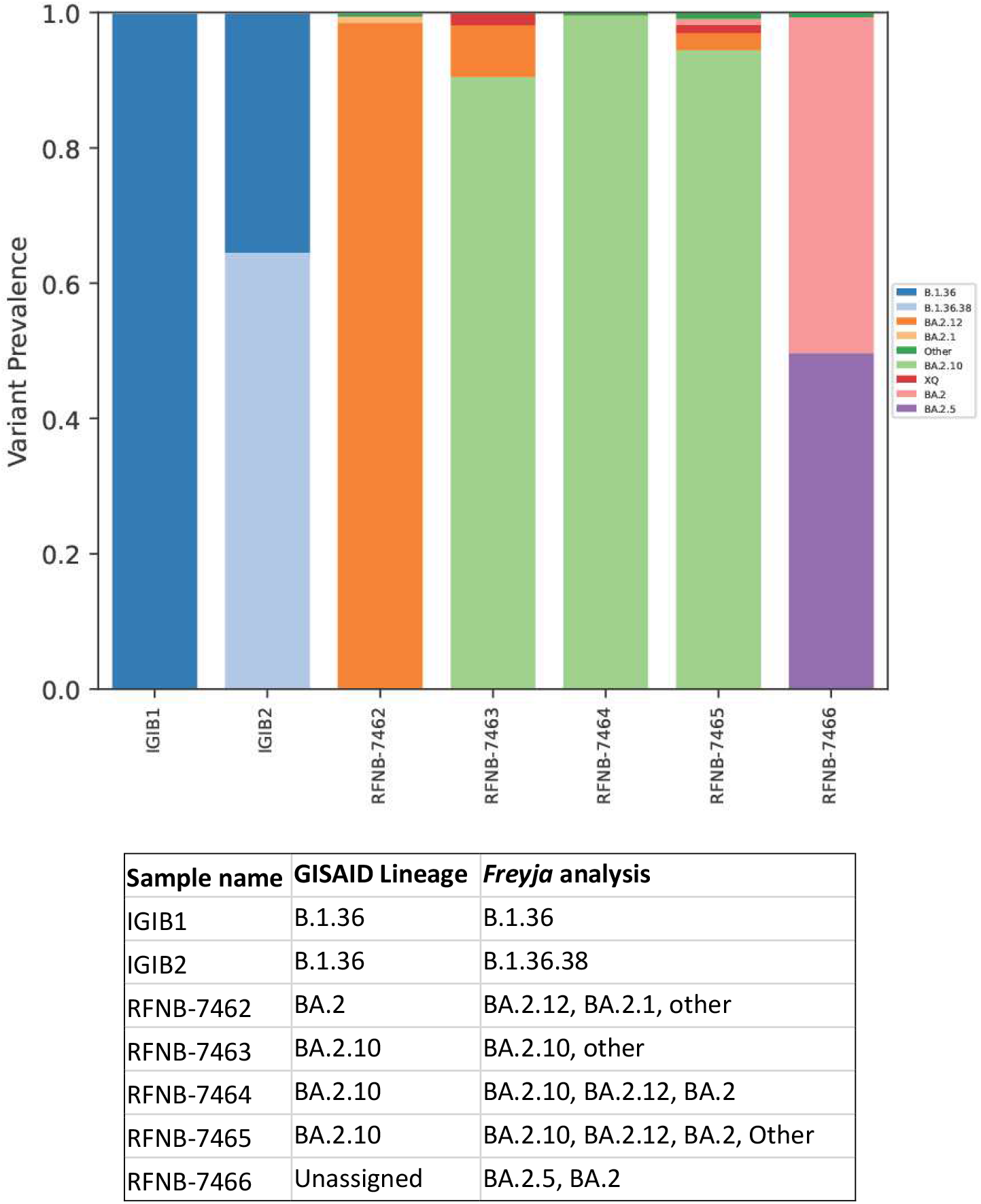
Sample deconvolution recovers relative virus abundance in clinical samples from Bangalore and Delhi.

**Fig. S4.**
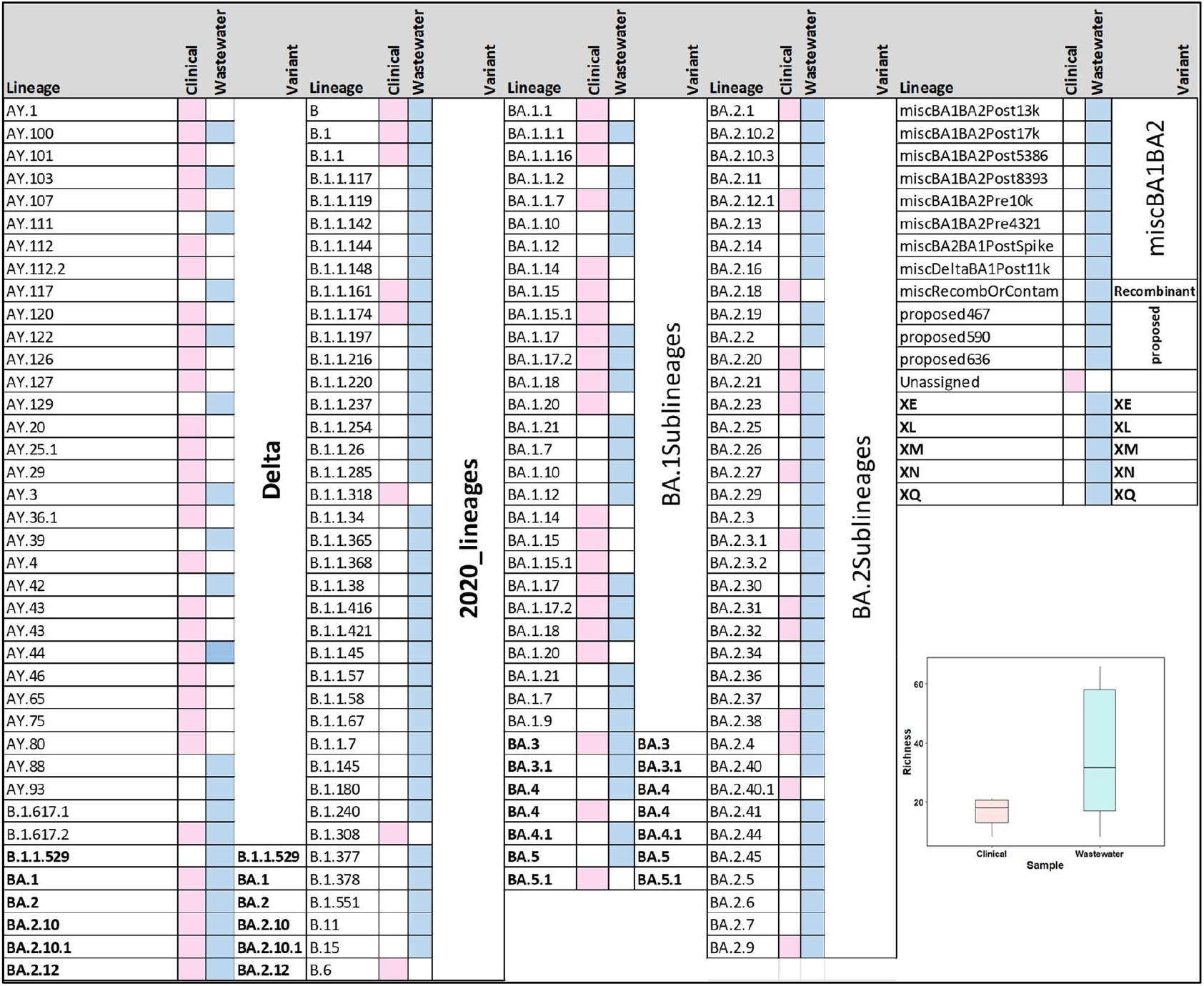
Comparison of SARS-CoV-2 lineage richness in Wastewater and Clinical samples.

**Fig. S5.**
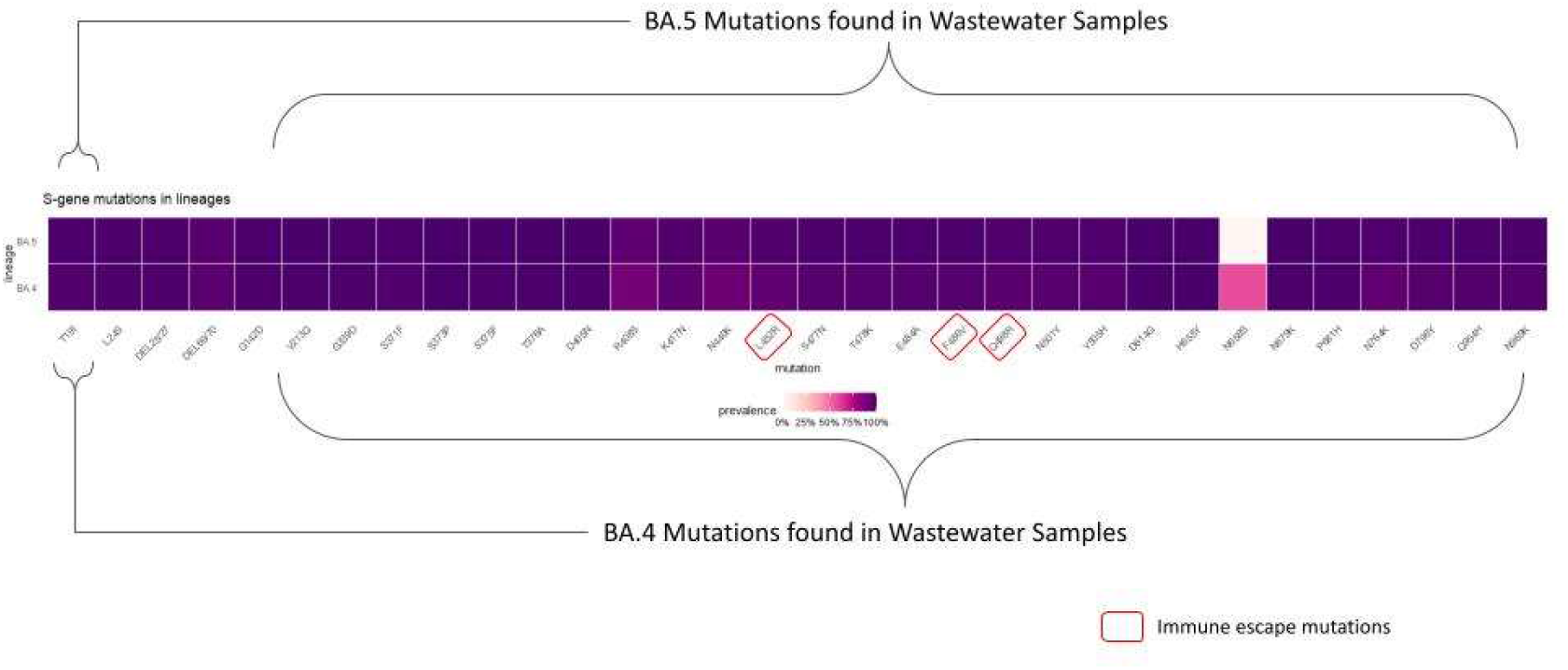
Spike gene mutations in BA.4 and BA.5 in wastewater samples

## References

1. COVIDPoops19, 2022. Summary of global SARS-CoV-2 wastewater monitoring efforts by UC Merced researchers [WWW Document]. Univ. Calif. Merced. URL 596 https://ucmerced.maps.arcgis.com/apps/dashboards/c778145ea5bb4daeb58d31afee389082 (Accessed 1.7.22).

2. Naughton, C.C., Roman, F.A., Grace Alvarado, A.F., Tariqi, A.Q., Deeming, M.A., Bibby, K., Bivins, A., Rose, J.B., Medema, G., Ahmed, W., Katsivelis, P., Allan, V., Sinclair, R., Zhang, Y., Kinyua, M.N., Author cnaughton, C., 2021. Show us the Data: Global COVID-19 Wastewater Monitoring Efforts, Equity, and Gaps. medRxiv 712 2021.03.14.21253564. https://doi.org/10.1101/2021.03.14.21253564

3. Aguiar-Oliveira et al. (2020). Wastewater-Based Epidemiology (WBE) and Viral Detection in Polluted Surface Water: A Valuable Tool for COVID-19 Surveillance-A Brief Review. Int J Environ Res Public Health. 10;17:9251.

4. Caicedo-Ochoa et al. (2020) Effective reproductive number estimation for initial stage of COVID-19 pandemic in Latin American countries. Int. J. Infect. Dis. 95, 316–318.

5. Crits-Christoph et al. (2021) Genome sequencing of Sewage Detects Regionally Prevalent SARS-CoV-2 Variants. Mbio. Vol. 12, No.1 e02703–2

6. Edition 4th (2011) Guidelines for drinking-water quality WHO chronicle, 38, pp. 104–108

7. Deshpande et al. (2003) Environmental surveillance system to track wild poliovirus transmission. Appl Environ Microbiol. 69, 2919–2927.

8. Hovi et al. (2001). Poliovirus surveillance by examining sewage specimens. Quantitative recovery of virus after introduction into sewerage at remote upstream location. Epidemiol. Infect. 127, 101–106.

9. Asghar et al. (2014) Environmental surveillance for polioviruses in the Global Polio Eradication Initiative. J Infect Dis. 210 (Suppl. 1): S294–303.

10. Ahmed et al. (2020). First confirmed detection of SARS-CoV-2 in untreated wastewater in Australia: A proof of concept for the wastewater surveillance of COVID-19 in the community. Sci. Total Environ., 728: 138764

11. Medema G, Been F, Heijnen L, et al. (2020) Implementation of environmental surveillance for SARS-CoV-2 virus to support public health decisions: Opportunities and challenges. Curr Opin Environ Sci Health, 17:49–71.

12. Peccia J, Zulli A, Brackney DE, et al. (2020) Measurement of SARS-CoV-2 RNA in wastewater tracks community infection dynamics. Nat Biotechnol, 38:1164–7.

13. La Rosa G, Iaconelli M, Mancini P, et al. (2020) First detection of SARS-CoV-2 in untreated wastewaters in Italy. Sci Total Environ, 736:139652.

14. Rambaut, A. et al. A dynamic nomenclature proposal for SARS-CoV-2 lineages to assist genomic epidemiology. Nat Microbiol 5, 1403–1407 (2020).

15. Turakhia, Y. et al. Ultrafast Sample placement on Existing tRees (UShER) enables real-time phylogenetics for the SARS-CoV-2 pandemic. Nat. Genet. 53, 809–816 (2021).

16. Wu, F., Xiao, A., Zhang, J., et al. SARS-CoV-2 titers in wastewater are higher than expected from clinically confirmed cases. mSystems, 5 (2020), 10.1128/mSystems.00614-20

17. Randazzo, W. et al. SARS-CoV-2 RNA in wastewater anticipated COVID-19 occurrence in a low prevalence area. Water Res. 181, 115942 (2020).

18. Brito AF, Semenova E, Dudas G, Hassler GW, Kalinich CC, Kraemer MUG, Ho J, Tegally H, Githinji G, Agoti CN, Matkin LE, Whittaker C; Danish Covid-19 Genome Consortium; COVID-19 Impact Project; Network for Genomic Surveillance in South Africa (NGS-SA); GISAID core curation team, Howden BP, Sintchenko V, Zuckerman NS, Mor O, Blankenship HM, de Oliveira T, Lin RTP, Siqueira MM, Resende PC, Vasconcelos ATR, Spilki FR, Aguiar RS, Alexiev I, Ivanov IN, Philipova I, Carrington CVF, Sahadeo NSD, Gurry C, Maurer-Stroh S, Naidoo D, von Eije KJ, Perkins MD, van Kerkhove M, Hill SC, Sabino EC, Pybus OG, Dye C, Bhatt S, Flaxman S, Suchard MA, Grubaugh ND, Baele G, Faria NR. Global disparities in SARS-CoV-2 genomic surveillance. medRxiv [Preprint]. 2021 Dec 9:2021.08.21.21262393. doi: 10.1101/2021.08.21.21262393. PMID: 34462754; PMCID: PMC8404891.

19. Karthikeyan et al. 2022 Wastewater sequencing reveals early cryptic SARS-CoV-2 variant transmission. Nature

20. Singanayagam, A. et al. Community transmission and viral load kinetics of the SARS-CoV-2 delta (B.1.617.2) variant in vaccinated and unvaccinated individuals in the UK: a prospective, longitudinal, cohort study. Lancet Infect. Dis. 22, 183–195 (2022).

20. Sapula SA, Whittall JJ, Pandopulos AJ, Gerber C, Venter H (2021) An optimized and robust PEG precipitation method for detection of SARS-CoV-2 in wastewater. Sci. Total Environ., 785: 147270.

21. Zdenkova K, Bartackova J, Cermakova E, Demnerova K, Dostalkova A, Janda V, Novakova Z et al. (2021) Monitoring COVID-19 spread in Prague local neighborhoods based on the presence of SARS-CoV-2 RNA in wastewater collected throughout the sewer network. medRxiv 2021.07.28.21261272

22. Zhang, T., Breitbart, M., W.H. Lee W.H. et al. RNA viral Community in Human Feces: prevalence of plant pathogenic viruses PLoS Biol., 4 (2005), Article e3, 10.1371/journal.pbio.0040003

23. Isaksoon, F. Lundy, L., Hedström, A., Székely, A.J., Mohamed, N. et al. Evaluating the use of alternative normalization approaches on SARS-CoV-2 concentrations in wastewater: Experiences from two catchments in Northern Sweden. Environments 2022, 9, 39. https://doi.org/10.3390/environments9030039

24. Singh, B. P., Madhusudan, J. V., Tiwari, A. K., Singh, S., & Das, U. D. (2020). Evaluation of EWMA control charts for monitoring spread of transformed observations of COVID-19 in India. Asian Journal of Research in Infectious Diseases, 5(2), 25–36.

25. Scrucca, L. (2004). qcc: an R package for quality control charting and statistical process control. dim (pistonrings), 1(200), 3.

26. Elias, C., Sekri, A., Leblanc, P., Cucherat, M. & Vanhems, P. The incubation period of COVID-19: A meta-analysis. Int. J. Infect. Dis. 104, 708–710 (2021).

27. Du, Z. et al. Shorter serial intervals and incubation periods in SARS-CoV-2 variants than the SARS-CoV-2 ancestral strain. J. Travel Med. 5, 1–3 (2022).

28. Grant, R. et al. Impact of SARS-CoV-2 Delta variant on incubation, transmission settings and vaccine effectiveness: Results from a nationwide case-control study in France. Lancet Reg. Heal. - Eur. 13, 1–12448 (2022)

29. Boucau et al. (2022) Duration of Shedding of Culturable Virus in SARS-CoV-2 Omicron (BA.1) Infection. DOI: 10.1056/NEJMc2202092

30. Wölfel, R, Corman, V.M., Guggemos, W, Seilmaier, M., Zange, S. Müller, et al. Virological assessment of hospitalized patients with COVID-2019 Nature, 1–10 (2020), 10.1038/s41586-020-2196-x

31. Rose, C., Parker, A., Jefferson, B., Cartmell, E., 2015. The characterization of feces and urine: a review of the literature to inform advanced treatment technology. Crit. Rev. Environ. Sci. Technol. 45 (17), 1827–1879.

32. Foladori P, Cutrupi F, Segata N, Manara S, Pinto F, Malpei F, Bruni L, La Rosa G. SARS-CoV-2 from faeces to wastewater treatment: What do we know? A review. Sci Total Environ. 2020 Nov 15;743:140444. doi: 10.1016/j.scitotenv.2020.140444.

33. Li, Heng (2014): Aligning sequence reads, clone sequences and assembly con*gs with BWA-MEM. figshare. Poster. https://doi.org/10.6084/m9.figshare.963153.v1

34. Li H, Handsaker B, Wysoker A, Fennell T, Ruan J, Homer N, Marth G, Abecasis G, Durbin R, and 1000 Genome Project Data Processing Subgroup, The Sequence alignment/map (SAM) format and SAMtools, Bioinformatics (2009) 25(16) 2078–9

35. Grubaugh, N. D. et al. An amplicon-based sequencing framework for accurately measuring intrahost virus diversity using PrimalSeq and iVar. Genome Biol. 20, 8 (2019)

36. Bhoyar RC, Jain A, Sehgal P, Divakar MK, Sharma D, Imran M, et al. (2021) High throughput detection and genetic epidemiology of SARS-CoV-2 using COVIDSeq next-generation sequencing. PLoS ONE 16(2): e0247115.

37. Julia L. Mullen, Ginger Tsueng, Alaa Abdel Latif, Manar Alkuzweny, Marco Cano, Emily Haag, Jerry Zhou, Mark Zeller, Emory Hufbauer, Nate Matteson, Kristian G. Andersen, Chunlei Wu, Andrew I. Su, Karthik Gangavarapu, Laura D. Hughes, and the Center for Viral Systems Biology. outbreak.info. outbreak.info https://outbreak.info/ (2021).

38. Oksanen J, Blanchet FG, Friendly M, Kindt R, Legendre P, McGlinn D, Minchin PR, O’Hara RB, Simpson GL, Solymos P, Stevens MHH, Szoecs E, Wagner H. 2017 vegan: community ecology package. https://github.com/vegandevs/vegan.

39. Kuznetsova, A., Brockhoff, P. B., & Christensen, R. H. B. (2017). lmerTest Package: Tests in Linear Mixed Effects Models. Journal of Statistical Software, 82(13), 1–26. https://doi.org/10.18637/jss.v082.i13

